# Mendelian Randomization analysis of the causal impact of body mass index and waist-hip ratio on rates of hospital admission

**DOI:** 10.1101/2020.07.14.20153742

**Authors:** Audinga-Dea Hazewinkel, Rebecca C. Richmond, Kaitlin H. Wade, Padraig Dixon

## Abstract

We analyze how measures of adiposity – body mass index (BMI) and waist-hip ratio (WHR) – causally influence rates of hospital admission. Conventional analyses of this relationship are susceptible to omitted variable bias from variables that jointly influence both hospital admission and adipose status. We implement a novel quasi-Poisson instrumental variable modelsin a Mendelian Randomization framework, identifying causal effects from random perturbations to germline genetic variation. We estimate the individual and joint effects of BMI, WHR, and WHR adjusted for BMI. We also implement multivariable instrumental variable methods in which the causal effect of one exposure is estimated conditionally on the causal effect of another exposure. Data on 310,471 participants and over 550,000 inpatient admissions in the UK Biobank were used to perform one-sample and two-sample Mendelian Randomization analyses. The results supported a causal role of adiposity on hospital admissions, with consistency across all estimates and sensitivity analyses. Point estimates were generally larger than estimates from comparable observational specifications. We observe an attenuation of the BMI effect when adjusting for WHR in the multivariable Mendelian Randomization analyses, suggesting that an adverse fat distribution, rather than a higher BMI itself, may drive the relationship between adiposity and risk of hospital admission.

## Introduction

Individuals with higher adiposity, as indexed by measures such as body mass index (BMI) and waist hip ratio (WHR), attend hospital more frequently than others (Buys et al., 2014; Chen, Jiang, & Mao, 2007; Han et al., 2009; Korda et al., 2015; Migliore et al., 2013; O’Halloran, 2020; Reeves, Balkwill, Cairns, Green, & Beral, 2014). Establishing the causal impact of adiposity on hospital admissions is an important step in understanding the impacts of adverse weight profiles on the health system. This importance stems from a number of considerations.

In the first instance, BMI (a marker of overall body fat) and WHR (a marker of regional adiposity) have been shown to be associated with increased incidence of various diseases (Corbin et al., 2016; Dale et al., 2017; Dalton et al., 2003; Folsom et al., 2000; Hu et al., 2007; Lyall et al., 2017; Staiano et al., 2012; Timpson et al., 2009) and all-cause and cause-specific mortality (Srikanthan, Seeman, & Karlamangla, 2009; Staiano et al., 2012; Wade, Carslake, Sattar, Davey Smith, & Timpson, 2018). Moreover, the incidence of adverse adiposity profiles is also increasing across the world. World Health Organization statistics identified 39% of men and 40% of women as overweight (BMI>25kg/m^2^) and 11% of men and 15% of women as obese (BMI>30kg/m^2^) worldwide (World Health Organization, 2016) in 2016. A positive association between BMI and healthcare costs has also been demonstrated (J. Cawley, 2015a; P. Dixon, Davey Smith, & Hollingworth, 2019; P. Dixon, Hollingworth, Harrison, Davies, & Davey Smith, 2020; Finkelstein, 2011; Kent, Fusco, et al., 2017; Kent, Green, et al., 2017; Withrow & Alter, 2011).

However, observational assessments of the association between adiposity and hospital attendance are challenged by endogeneity attributable to unobserved confounding and reverse causation, precluding accurate causal inference (Auld & Grootendorst, 2011; J. Cawley, 2015a, 2015b; John Cawley & Meyerhoefer, 2012)

In this paper, we introduce the first Mendelian Randomization analysis to use hospital admissions as an outcome. We used UK Biobank data from over 300,000 adults aged 39-72, with over 550,000 in-patient hospital admissions in relation to three three related exposures: BMI, WHR and WHR adjusted for BMI (WHRadjBMI).

Mendelian randomization is an instrumental variable approach that allows for the robust estimation of the causal effect of an exposure or treatment variable (e.g., BMI) on an outcome (e.g., hospital admissions) (G. Davey Smith & Hemani, 2014; Haycock et al., 2016). The identifying assumption of Mendelian Randomization is the quasi-random perturbation of germline genetic variation that occurs at conception. Elements of this variation are known to associate with traits such as BMI and waist-hip ratio, and may be used as instrumental variables in causal analysis relating the effects of these adiposity-related exposure variables to hospital admission outcomes.

Mendelian Randomization reduces or eliminates problems of confounding and other types of bias common to conventional studies of the associations between measures of adiposity and healthcare-related resource use. For example, Mendelian Randomization can rule out reverse causation, since germline variants are determined at conception, and, in principle, should not be affected by confounding from unmeasured variables, given the quasi-random allocation of variants from parents to offspring at conception.

We implement three broad classes of estimator to assess these causal associations. We first implemented a generalized linear model (GLM) version of the familiar two-stage least squares (2SLS) estimator. Our outcome was hospital admission counts for subjects observed for varying lengths of time, reflecting the duration of follow-up available in our outcomes data accounting for time of recruitment and censoring due to end of follow-up or death. As Poisson regression models are linear on the logarithmic scale, the second stage of the standard 2SLS instrumental variable estimator was replaced by a Poisson regression. These models were just identified, utilizing a genetic risk score (a single summary measure indicating genetic liability to the exposure of interest) as a single instrumental variable.

Valid instrumental variables are associated with the exposure of interest, are conditionally independent of known and unknown omitted variables, and affect the outcome only via their effect on the exposure (the exclusion restriction). The most likely source of violation of these assumptions in Mendelian Randomization is the exclusion restriction, and for that a reason a variety of estimators were employed to test the robustness of our results to the presence and consequences of any violations of this assumption. The second broad class of estimators therefore involved over-identified models that allow that exclusion restriction to be relaxed for some or all variants, at the cost of other assumptions that we set out below.

Finally, in addition to testing the direct, individual effect of these exposures on admissions, we also implemented multivariable instrumental variable models, in which the causal effect on one exposure is estimated conditional on the causal effect of another exposure. This permits evaluation of whether the causal effect of one exposure on admissions is mediated by another exposure. For example, this approach allows for estimation of the direct effect of BMI on hospital admissions that is not mediated via the indirect effect of BMI on waist hip ratio, and (simultaneously) the direct of effect of waist hip ratio that is not mediated by the indirect effect of BMI on this outcome.

The rest of the paper is set out as follows. Below, we briefly introduce the idea of genetic variants as instrumental variables. We then introduce the data used for our analysis. We describe in detail our methods for estimating the direct and indirect effect of these exposures on admissions, before concluding with our interpretation of these results. Our results indicate that the effect of various measures of adiposity on admission may be larger than conventional observational analysis would indicate, and that regional adiposity (as indexed by WHR) may play a particularly important role in influencing the rate of hospital admissions.

### 1.1 Genetic variants as instrumental variables

Many introductions to MR are available both in general (George Davey Smith & Ebrahim, 2003; Haycock et al., 2016; Pingault et al., 2018) and in relation to health economic outcomes (P. Dixon et al., 2019; Padraig Dixon, Davey Smith, von Hinke, Davies, & Hollingworth, 2016; Harrison et al., 2020; von Hinke Kessler Scholder, Smith, Lawlor, Propper, & Windmeijer, 2011; Von Hinke, Davey Smith, Lawlor, Propper, & Windmeijer, 2016). Here we briefly review the instrumental variable assumptions in the context of Mendelian Randomization.

Certain parts of the genome are subject to variation between individuals in a population. At each of these points of variation, offspring inherit an allele – the specific form of genetic variation – from each of their parents according to Mendel’s first and second laws of inheritance. Mendel’s first law describes random segregation of alleles from parents to offspring. Mendel’s second law describes the independent assortment for different traits of these alleles. Together, the two laws imply that offspring have an equal chance of inheriting an allele from either parent and that these alleles are inherited independently from one another. The allocation of these variants is therefore random, conditional on parental genomes.

It is this form of conditionally random allocation and its use as an identification mechanism in instrumental variable analysis that is known as Mendelian Randomization (George Davey Smith & Ebrahim, 2003). Mendelian Randomization may therefore be interpreted as a type of natural experiment, in which individuals or groups of individuals are allocated to groups indicating higher or lower genetic liability to (for example) higher BMI or WHR. Under the assumptions of instrumental variable analysis, this quasi-random allocation can be used to make causal claims about the effect of these types of exposures on hospital admissions.

We study single nucleotide polymorphisms (SNPs), which are one form of genetic variation (amongst others) that are subject to inheritance under Mendel’s first and second laws. A SNP refers to single change in one of the nucleotides that make up the code of the genome. Nucleotides in DNA are in turn made up in part of the nucleobases (adenine (A), cytosine (C), guanine (G) or thymine (T) which comprise this code. A SNP will therefore involve a substation of one of these “letters” in the genetic code for another. The possible versions of the SNP at a specific point are the alleles for that location in the genome. Some SNPs are associated with the expression of particular traits or phenotypes, including several adiposity-related phenotypes such as BMI and WHR.

The association of SNPs with phenotypes, together with their conditionally random allocation from parents to offspring, indicate the potential for their analysis as instrumental variables. Humans are diploid, meaning that they have two copies of each chromosome. We may therefore treat SNPs as count variables – humans may have a SNP on both chromosomes (n=2), only on one (n=1) or on neither chromosome (n=0).

Valid instrument variables are associated with the exposure of interest, are independent of all confounding omitted variables (whether measured or unmeasured) and affect the outcome only via the exposure of interest. We briefly unpack these requirements in relation to Mendelian Randomization.

Relevance – the requirement that instruments are not independent of exposures – can be determined from genome wide association studies (GWASs), which trawl the genome for signals of association between a predefined exposure and regions of potential genetic variation (McCarthy et al., 2008). Replicated evidence of association from large, well-powered GWASs are the most robust means of establishing the relevance criterion for specific SNPs.

The second requirement, that of independence from confounding omitted variables, is sometimes interpreted as the requirement that the instrument be “as good as randomly assigned” (J. Angrist & Pischke, 2009). As genetic variation is determined at conception, it necessarily occurs before many later life circumstances and events such as socioeconomic status, education level, and the local environment. This ensures independence from most potential confounding variables.

Nevertheless, there are a few different means by which this assumption might be violated. Events that are connected with the time of conception, such as year of birth and sex, may confound this association. This requirement will also be violated if there are differences in subgroups defined by allele frequencies that also differ in disease or trait susceptibility. For example, allele frequencies differ by ancestry, and environments are not necessarily the same between groups of different ancestry. Allele subgroups may become correlated with the environment for other reasons. For instance, assortative mating describes the mating of genetically similar individuals. Over time, this will tend to lead to a non-random clustering of alleles, potentially violating this assumption.

The third requirement for instrumental variable analysis, the exclusion restriction, may be violated through two principal mechanisms in Mendelian Randomization analysis. The first is via so-called linkage disequilibrium, which refers to the fact that SNPs in close physical proximity tend to be inherited together. Use of one of these SNPs may therefore also reflect the effect of other SNPs not intended to be included in the analysis.

The second and more challenging potential violation of this assumption is via pleiotropy (Hemani, Bowden, & Davey Smith, 2018). Pleiotropy is the effect of a genetic variant on more than one phenotype. The exclusion restriction will be violated if a SNP associated with (for example) BMI also affects the outcome through a BMI-independent channel. This is known as horizontal pleiotropy (G. Davey Smith & Hemani, 2014). The exclusion restriction will not be violated if the other phenotype does not affect the outcome, or if the other phenotype is intermediate between the exposure of interest and the outcome (the latter is known as vertical pleiotropy).

Below, in the Methods section, we describe the ways in which we attempted to account for the requirements for valid instrumental variable analysis.

### Data

#### UK Biobank data

The UK Biobank study is a resource of phenotypic, genetic, electronic health record and death registry data, collected from over 500,000 individuals, from 2006 to 2010 (Bycroft et al., 2018; Collins, 2012; Sudlow et al., 2015). Participants were aged 39-72 years at recruitment and were predominantly of White British ethnicity (Sudlow et al., 2015). The UK Biobank received ethical approval from the North West-Haydock Research Ethics Committee (reference 11/NW/0382).

For 465,373 participants, information on hospital inpatient admissions was available through linked Hospital Episode Statistics (HES) data. Inpatient admisisons involve a patient occupying a bed for some period, but does not necessarily imply an overnight hospital stay. From this set, 310,471 participants were considered eligible for analysis (Figure 1). Participants were removed when admission information was incorrect, when the genetic data did not meet the standard of a documented in-house quality control procedure (Mitchell, 2019), and when BMI and/or WHR measurements were absent. All analyses (both observational and instrumental variable models) were restricted to individuals of White British ancestry to avoid confounding by differential ancestry under the second instrumental variable assumption. Participants matching one or more of the exclusion criteria were removed (N=154,902) (Figure 1).

**Figure 1.**
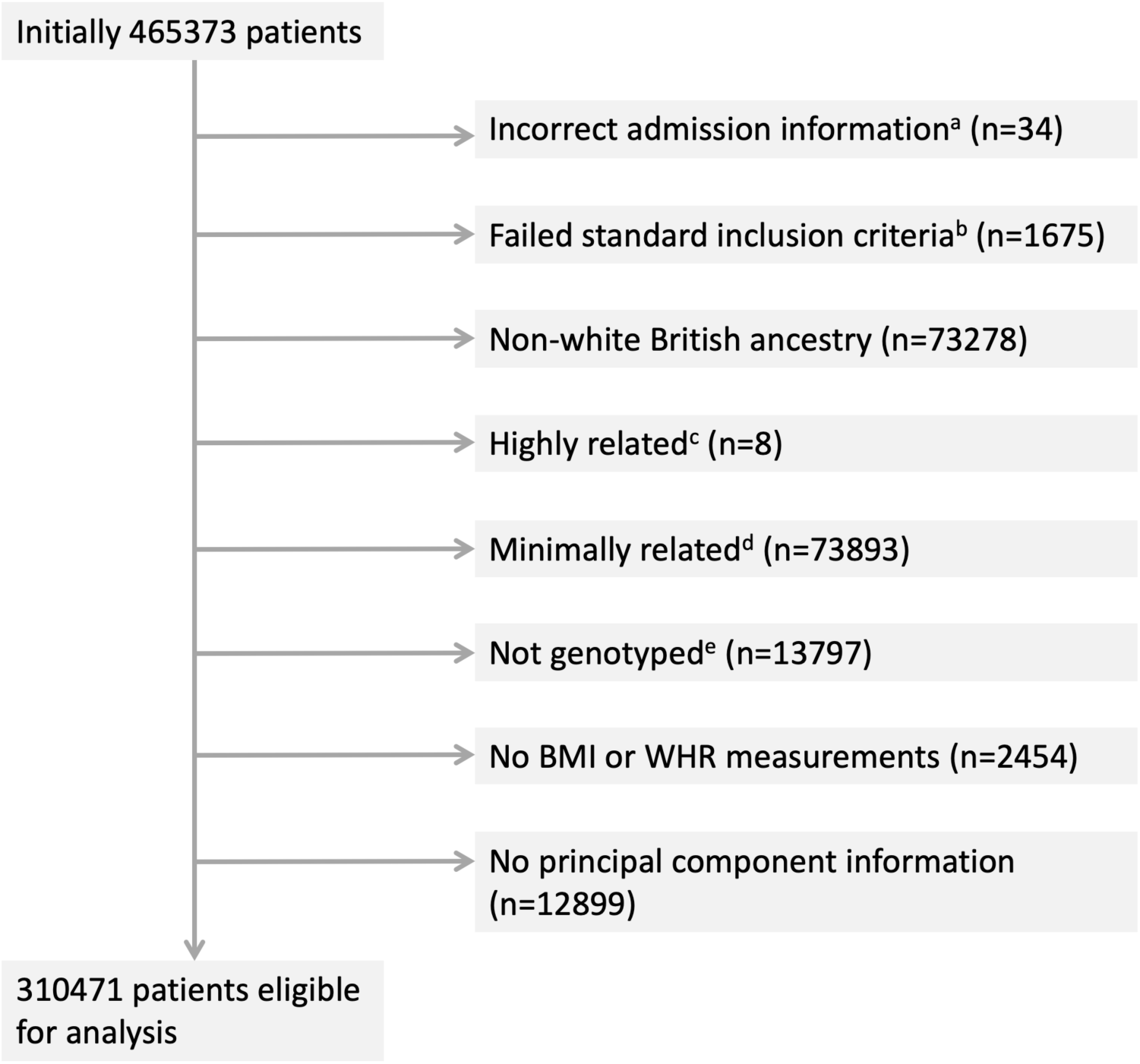
Participant inclusion diagram. **NOTES TO FIGURE**: Eight exclusion categories are shown with the corresponding numbers of participants per category. On removal of duplicates a total of 154902 unique participants are considered ineligible for analysis. a) admissions prior to study start date, post death/censoring date or registered death prior to study start; b) individuals that have a mismatch between genetically inferred and reported gender, individuals with sex chromosome types putatively different from XX or XY and individuals that are outliers in heterozygosity and missing rate; c) individuals related to more than 200 other participants; d) on exclusion a maximal set of unrelated individuals is retained; e) not genotyped for the exposures of interest (BMI, WHR, WHRadjBMI)

#### Adiposity measures

Measures on weight, bio-impedance, height, waist circumference and hip circumference were collected at the baseline UK Biobank assessment. Weight and bio-impedance were measured using the Tanita BC-418MA body composition analyzer. Standing height was measured using a Seca 202 height measure. BMI was calculated as weight divided by height squared (kg/m^2^), and using electrical impedance. When the first measure was unavailable, values were supplemented with the latter. WHR was calculated by dividing waist circumference by hip circumference, measured with a Wessex non-stretchable spurring tape measure (UK Biobank, 2011).

#### Hospital admission counts

Hospital admission count was derived using electronic health records (“Hospital Episode Statistics”) linked to the UK Biobank study. For a given hospital admission, a patient may have multiple “episodes” of care. Admissions were therefore defined where an individual had episodes starting on separate dates, excluding incomplete episodes, episodes with inconsistent or overlapping start and end dates and accounting for patient transfers. Code to define the admissions variable is available from https://github.com/pdixon-econ/admissions-biobank.

#### Genetic variants

Estimates for 77 genetic variants associated with BMI at a genome-wide significance level (p<5×10^−8^) in the largest genome-wide association study meta-analysis of a combined number of up to 322,154 individuals of European descent (not including UK Biobank) were obtained from the Genetic Investigation of Anthropometric Traits (GIANT) consortium (Locke et al., 2015). Individual-level genetic data of sufficient quality (Mitchell, 2019) ^34^ was available from UK Biobank for 76 of these 77 SNPs. Genetic variants associated with WHR and WHRadjBMI were also obtained from the GIANT consortium, with 39 and 48 SNPs, respectively, identified in relation to WHR at p<5×10^−8^, in a meta-analysis of up to 210,088 individuals (Shungin et al., 2015).

## Methods

This section describes our approach to modelling the association between healthcare costs and the three measures of adiposity. We implement both conventional observational models and instrumental variable models. Both types of model implement versions of Poisson regression.

### Poisson modelling of admissions data

A Poisson regression model applies a generalized linear model with a logarithmic link function under the assumption that the response variable is Poisson distributed and that the logarithm of the expected value *μ* can be expressed in a linear combination of *k* parameters (see Formula 1 below).

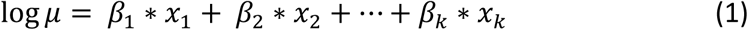

UK Biobank participants were recruited between 2006 and 2010 and for each participant hospital admissions were counted from recruitment to study censoring with the latter given by either death or 31 March 2015, the date at which the linked Hospital Episode Statistic data were censored for this analysis. We ignore emigration, which is estimated to occur at low rates (0.3%) in this cohort (Fry et al., 2017).

To correct for the varying times on study, the logarithm of observed person-years *t* is added to the linear equation as an offset (Formula 2).

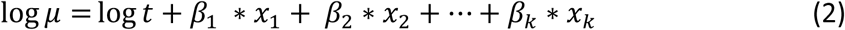

The modeled outcome can now be interpreted as a rate rather than a count, as becomes apparent when restructuring equation (2) to equation (3).

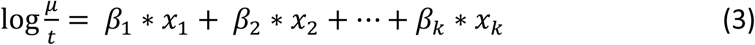

Let *x*_*1*_ and *β*_1_ represent the exposure BMI and the corresponding parameter estimated from the Poisson regression. Then with *t* in years and BMI in kg/m^2^the exponent of the coefficient exp(*β*_1_) is the factor by which the mean value of the outcome is multiplied for a 1 kg/m^2^ increase in BMI. More generally, with the remaining variables are held constant, the yearly hospital admission rate increases with a factor *exp(β*_*1*_)^*n*^ for every *n* unit increase in BMI. Values of the coefficient >1 indicate an increase in admission rate and values <1 indicate a decrease.

### Multivariable observational analyses

Observational estimates were obtained by regressing each exposure on the outcome in a conventional (i.e. without instrumental variables) Poisson regression, with time in UK Biobank as the offset. Unadjusted estimates were obtained alongside estimates adjusted for age at study entry, sex, and a number of categorical variables. To make full use of the available data, we imputed missing values in a 10-fold imputation approach and pooled the resulting coefficients and standard errors using Rubin’s rules (Rubin, 1996).

### Instrumental variable Poisson models

Mendelian Randomization methods are traditionally applied to continuous or binary outcome data, whereas count data such as hospital admission data is frequently modeled using Poisson regression. Here, the outcome was given by hospital admission counts for subjects that were observed for varying lengths of time. As the Poisson model is linear on the logarithmic scale, the second stage of the Mendelian Randomization regression that estimates the gene-outcome association can be replaced by a Poisson regression.

We conducted both one- and two-sample Mendelian Randomization analyses. In a one-sample framework, a single sample of individual-level genetic and phenotypic data is used to obtain estimates of both the gene-exposure association (*β*_*exp*_) and the gene-outcome association (*β*_*out*_). External weightings from the GIANT consortium were used for the gene-exposure associations.

The ratio of the coefficients (the Wald ratio) gives the causal 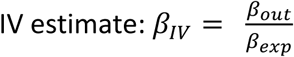. When a single instrument is used in a linear model, the Wald ratio is identical to the 2SLS estimator. Here, we implement a generalized linear model version of this estimator to allow for a Poisson model in the second stage.

Note that the numerator and the denominator in the Wald ratio need not come from the same sample (J. D. Angrist & Krueger, 1992). In a two-sample Mendelian Randomization framework, the exposure and outcome coefficients are obtained from separate, independent samples from similar populations (G. Davey Smith & Hemani, 2014; Haycock et al., 2016). This approach may offer better efficiency than a one-sample approach if larger sample sizes are available when using data from more than one sample. A further important advantage is that the two-sample approach particularly facilitates methods to test the sensitivity of results to possible violations of the exclusion restriction, as we discuss below.

We therefore implemented both one- and two-sample Mendelian Randomization models. For both the one- and two-sample analyses we implemented a model where the second stage linear regression of the outcome *Y*_*adm*_ on the respective SNPs is replaced by a Poisson regression with the person years on study *t* as offset. Let *β*_exp_ be the exposure coefficient obtained from a linear regression of the exposure *Y*_*exp*_ on the genetic instrument or risk score *G* (see Formula 4), and *β*_out_ the outcome coefficient on the logarithmic scale obtained from a Poisson regression of the outcome *Y*_*out*_ on the genetic instrument *G* (see Formula 5).

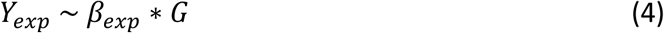

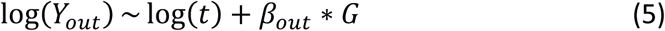

As the Poisson model is linear on the log scale, the two coefficients are compatible and a valid ratio can be obtained, with the final IV estimate *β*_*IV*_ of the rate coefficient given by the exponent of this ratio (6).

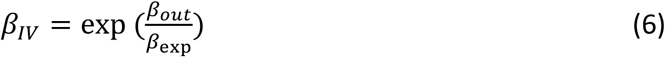

The coefficient *β*_*IV*_ is the factor by which the yearly hospital admission rate increases for each unit of exposure, again with values >1 indicating an increase in admission rate and values <1 indicating a decrease.

The Poisson model assumes an outcome distribution such that the outcome mean is equal to the outcome variance. As the hospital admission count variance (55.3) was greater than the mean hospital admission count (1.89), we instead used a quasi-Poisson model (Ver Hoef & Boveng, 2007). Standard errors for the causal IV estimates were estimated using Taylor series expansions (Thomas, Lawlor, & Thompson, 2007).

### One-sample just-identified Mendelian randomization

Weighted genetic risk scores (GRS)s were constructed for BMI (76 SNPs) (Locke et al., 2015), WHR (39 SNPs) (Shungin et al., 2015) and WHRadjBMI (48 SNPs) (Shungin et al., 2015). To ensure a meaningful interpretation of the score, the exposure-increasing allele for each genetic variant was chosen as the effect allele (Wade et al., 2018). For each study participant, the dosage for each relevant genetic variant was extracted from the UK Biobank genetic data and weighted with the effect size reported by the GIANT consortium. Following this, the weighted dosages were summed and divided by the sum of all effect sizes, giving a single GRS for each exposure representing an estimate for the average number of exposure-increasing alleles. Estimates were adjusted for age, sex and the first 40 genetic principal components (PCAs) to comply with the second instrumental variable assumption of conditional independence (“as good as randomly assigned”).

To estimate the effect of WHRadjBMI on hospital admissions, we generated the residuals of a regression of WHR on BMI as an exposure, which gave an estimate for the predictive performance of the WHR component that cannot be linearly predicted by BMI.

We also considered the effects of BMI and WHR when estimated in a joint model, using a multivariable Mendelian Randomization approach. Multivariable Mendelian Randomization aims to estimate the causal effect of multiple exposures simultaneously. In contrast to univariable Mendelian Randomization, which estimates the total effect of an exposure on the outcome, Multivariable Mendelian Randomization estimates the direct effect of each exposure conditioning on the causal effects of the SNPs on the other exposure (Sanderson, Davey Smith, Windmeijer, & Bowden, 2019). We regressed BMI and WHR separately on the full combined set of SNPs and regressed the fitted values of both on hospital admission count in a Poisson regression with time as the offset. Standard errors were obtained through a 10,000-fold full-sample bootstrap (Efron & Tibshirani, 1993).

For BMI, estimates of hospital admission rates per year were obtained per BMI unit (1 kg/m^2^) and per BMI standard deviation (SD). For WHR, WHRadjBMI, and the WHR residuals, estimates were obtained per 0.10 WHR unit and per WHR SD. The relevant SDs were calculated directly from the UK Biobank data, yielding SDs of 4.74 kg/m^2^ and 0.09 for BMI and WHR, respectively.

### Two-sample over-identified summary Mendelian randomization

We employed a variety of over-identified methods to assess the causal association between adiposity and hospital admissions in two-sample Mendelian Randomization analysis. Four two-sample Mendelian Randomization approaches were used to investigate the effect of BMI, WHR and WHRadjBMI on yearly hospital admission rate: 1) the random effects exact weights inverse-variance weighted (IVW) estimator (Bowden et al., 2019); 2) the random effects Mendelian Randomization Egger estimator (Bowden, Davey Smith, & Burgess, 2015); 3) the penalized median estimator (Bowden, Davey Smith, Haycock, & Burgess, 2016); 4) the weighted mode estimator (Hemani, Bowden, et al., 2018).

All of these approaches make distinct assumptions about whether and how the exclusion restriction might be violated. Precise technical details are available in the respective references. Here, we provide an overview of these details and some intuition for their implementation.

A useful starting point is to approach this type of instrumental variable analysis from the perspective of meta-analysis (Bowden & Holmes, 2019). If each SNP is treated as the outcome of a natural experiment occurring at conception, then an overall effect estimate across many SNPs may be obtained by performing meta-analysis. For example, for the random effects models, Wald ratios are estimated for each SNP separately and combined using a random effects meta-analysis approach. The traditional IVW estimator uses weights derived from the inverse variance of the SNP-outcome coefficient. The exact weights IVW estimator derives weights in a slightly different way, using a limited information maximum-likelihood (LIML) approach in which the weight term is allowed to be a function of the causal-effect parameter. This approach therefore also ensures the estimator is naturally robust against regression-dilution bias (Bowden et al., 2019).

This approach assumes either that there is no horizontal pleiotropy in violation of the exclusion restriction, or that any horizontal pleiotropy balances out such that point estimates of effect are not biased. Violations of the exclusion restriction induced by horizontal pleiotropy may be apparent if the effect of a SNP or set of SNPs is large relative to the mean effect of all SNPs.

Heterogeneity of this type was assessed in two related ways. The first was via Cochran’s Q statistic, the two-sample analogue of the Sargan test for overidentification. Heterogeneity can be assessed by comparing the Q statistic (Formula 7) to the critical values of a chi-squared distribution:

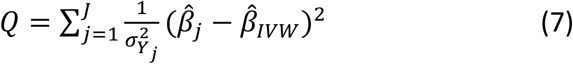

This assumes up to *J* SNPs; 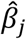 and 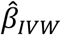measure respectively the effect estimates for SNP *j* and the overall inverse variance (IVW) weighted effect over all *J* SNPs. The variance of the SNP-outcome association is 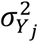.

The second, related means of assessing heterogeneity was based on Rucker’s Q’ statistic (Bowden, Hemani, & Davey Smith, 2018), which first requires a discussion of the Mendelian Randomization-Egger approach. The random effects IVW model constraints the overall IVW-regression line to pass through the origin; if this were not the case, then some or all variants would be violating the exclusion restriction since the causal effect (absent violations of the exclusion restriction) is determined only by the Wald ratio. The random effects Mendelian Randomization-Egger model does not constrain the intercept to zero. The Mendelian Randomization-Egger regression intercept can be interpreted as the average pleiotropic effect of all variants. Verifying whether the observed intercept is statistically different from zero serves as a test for horizontal pleiotropy (Bowden et al., 2015).

In essence, this amounts to testing and then conditioning on any horizontal pleiotropy, at the cost of further assumptions concerning the association between instrument strength and the direct pleiotropic effect of variants – on this assumption, see further detail in (Bowden et al., 2015; Stephen Burgess & Thompson, 2017). Rucker’s Q’ statistic may be calculated by estimating the Mendelian Randomization-Egger model, adjusting for any mean pleiotropic effect, and then testing if any residual heterogeneity is present.

The third estimator is the median estimator, which is given by the median ratio estimate of ordered Wald ratios. The intuition for this estimator is that a consistent estimate will be obtained from the median estimate if at least 50% of the SNPs are valid; invalid estimates will contribute no weight to the overall estimate provided that at least this proportion of SNPs are valid. The penalized weighted median estimator will be consistent when at least 50% of the weights come from valid instruments. We report results from the penalized weighted median estimator, in which Cochran’s Q statistic is used to quantify heterogeneity and more heterogeneous, outlying variants are down-weighted (Bowden et al., 2016).

The mode estimate is given by the mode of the Wald ratio estimates and will consistently estimate the causal effect even if more than half of the SNPs are invalid, provided the largest homogeneous cluster of SNPs is valid. Both the weighted median and mode estimators use weights derived from the inverse variance of the ratios of the gene-outcome and gene-exposure association estimates and are, under different assumptions, robust to outliers and invalid instruments (Hemani, Bowden, et al., 2018).

Gene-exposure association coefficients for the two-sample Mendelian Randomization analyses were obtained from the GIANT consortium (Locke et al., 2015; Shungin et al., 2015) and gene-outcome association coefficients from the UK Biobank data (Bycroft et al., 2018; Collins, 2012; Sudlow et al., 2015).

Two-sample Mendelian Randomization estimates were obtained for BMI, WHR and WHRadjBMI individually and for BMI and WHR jointly in a multivariable two-sample Mendelian Randomization IVW analysis (Sanderson et al., 2019). For BMI, estimates of hospital admission rates per year were obtained per BMI unit (1 kg/m^2^) and SD. For WHR and WHRadjBMI estimates were obtained per 0.10 WHR unit and SD. SDs were calculated by taking the median SD across all studies used to obtain the summary measures, giving SDs of 4.60 kg/m^2^and 0.07 for BMI and WHR, respectively. Plots of the two Q (Cochran’s Q and Rucker’s Q’) statistics were calculated for each SNP in a leave-one-out analysis and were used for the visual identification of outliers, which were removed in a sensitivity analysis with the purpose of verifying estimator consistency.

A threshold of R^2^<0.001 to account for linkage disequilibrium (LD) was employed for the two-sample Mendelian Randomization analyses. Post LD-correction, 64, 34 and 45 SNPs were retained for BMI, WHR and WHRadjBMI, respectively. For the multivariable two-sample Mendelian Randomization analysis, 70 SNPS were retained after a joint LD adjustment for BMI and WHR.

All analyses were performed in R version 3.6.1 (R Development Core Team, 2014). R packages *TwoSampleMendelian Randomization* (Hemani, Zheng, et al., 2018), *RadialMendelian Randomization* (J. Bowden et al., 2018), and *MVMendelian Randomization* (Sanderson et al., 2019) were used for the two-sample summary Mendelian Randomization analyses and *Amelia* (Honaker, 2019) was used for the multiple imputation performed in the conventional multivariable analyses. An *R* code appendix documenting all analytical steps taken is provided in the supplementary materials.

## Results

This section sets out the results of the analysis described above. We begin with a summary of descriptive statistics and any missingness in these variables. We then introduce results from our observational analysis, which serve as a useful benchmark against which to judge the Mendelian Randomization instrumental variable models. We then present the results from the one-sample Mendelian Randomization using the GRS instruments, before considering the over-identified two-sample models that test potential violations of the exclusion restriction.

### Descriptive statistics

The 310,471 participants included in the analysis sample had an average age of 57.40 years (SD: 7.99), with a BMI of 27.38kg/m^2^ (SD: 4.74) and WHR of 0.87 (SD: 0.09). Of these participants, 53.66% were female. Average follow-up time was 6.05 years (SD: 0.91). BMI, WHR, sex and age distributions were comparable across the UK Biobank and the GIANT consortium populations (Bycroft et al., 2018; Locke et al., 2015; Shungin et al., 2015). Demographics for the UK Biobank participants are given in Table 1.

**Table 1.** Patient demographics at baseline for N=310471 patients. For continuous variables mean and standard deviation are given, for categorical variables counts and percentages per category.

| Characteristic | N / mean (SD) | % | Characteristic | N | % |
| --- | --- | --- | --- | --- | --- |
| Age at entry | 57.402 (7.988) | 100 | Qualifications |  |  |
| Missing | 0 | 0 | A levels/AS levels or equivalent | 35268 | 11.4 |
| Sex |  |  | College or University degree | 96670 | 31.1 |
| Female | 166610 | 53.7 | CSEs or equivalent | 17714 | 5.70 |
| Male | 143861 | 46.3 | NVQ or HND or HNC or equivalent | 20418 | 6.60 |
| Missing | 0 | 0 | O levels/GCSEs or equivalent | 69815 | 22.5 |
| BMI | 27.385 (4.743) | 100 | Other professional qualifications eg: nursing, teaching | 15821 | 5.10 |
| Missing | 0 | 0 | Missing | 54765 | 17.6 |
| WHR | 0.872 (0.09) | 100 | Employment |  |  |
| Missing | 0 | 0 | Doing unpaid or voluntary work | 1298 | 0.40 |
| Alcohol frequency |  |  | Full or part-time student | 544 | 0.20 |
| Daily or almost daily | 67391 | 21.7 | In paid employment or self-employed | 175679 | 56.6 |
| Never | 19900 | 6.4 | Looking after home and/or family | 8042 | 2.60 |
| Once or twice a week | 81161 | 26.1 | Retired | 109314 | 35.2 |
| One to three times a month | 34254 | 11.0 | Unable to work because of sickness or disability | 9043 | 2.90 |
| Special occasions only | 32382 | 10.4 | Unemployed | 4212 | 1.40 |
| Three or four times a week | 75173 | 24.2 | Missing | 2339 | 0.80 |
| Missing | 210 | 0.10 | Townsend deprivation |  |  |
| Days exercise |  |  | 1st Quint. (-6.258, -4.014] | 62055 | 20.0 |
| 0 days | 110190 | 35.5 | 2nd Quint. (-4.014, -2.945] | 62002 | 20.0 |
| 1 day | 42722 | 13.8 | 3rd Quint. (-2.045, -1.683] | 62028 | 20.0 |
| 2 days | 47400 | 15.3 | 4th Quint. (-1.683, 0.709] | 62030 | 20.0 |
| 3 days | 40976 | 13.2 | 5th Quint. (0.709, 10.588] | 62016 | 20.0 |
| 4 days | 19061 | 6.10 | Missing | 341 | 0.10 |
| 5 days | 19800 | 6.40 |  |  |  |
| 6 days | 5758 | 1.90 |  |  |  |
| 7 days | 10315 | 3.30 |  |  |  |
| Missing | 14249 | 4.60 |  |  |  |

BMI = body mass index, SD = standard deviation, WHR = waist-hip-ratio

A total of 588,147 in-patient hospital admissions was recorded, with 79% of participants admitted twice or less, and 47% without a single admission. Table 2 gives an overview of hospital admission counts across BMI categories, WHR quantiles, age quantiles and gender.

**Table 2.**
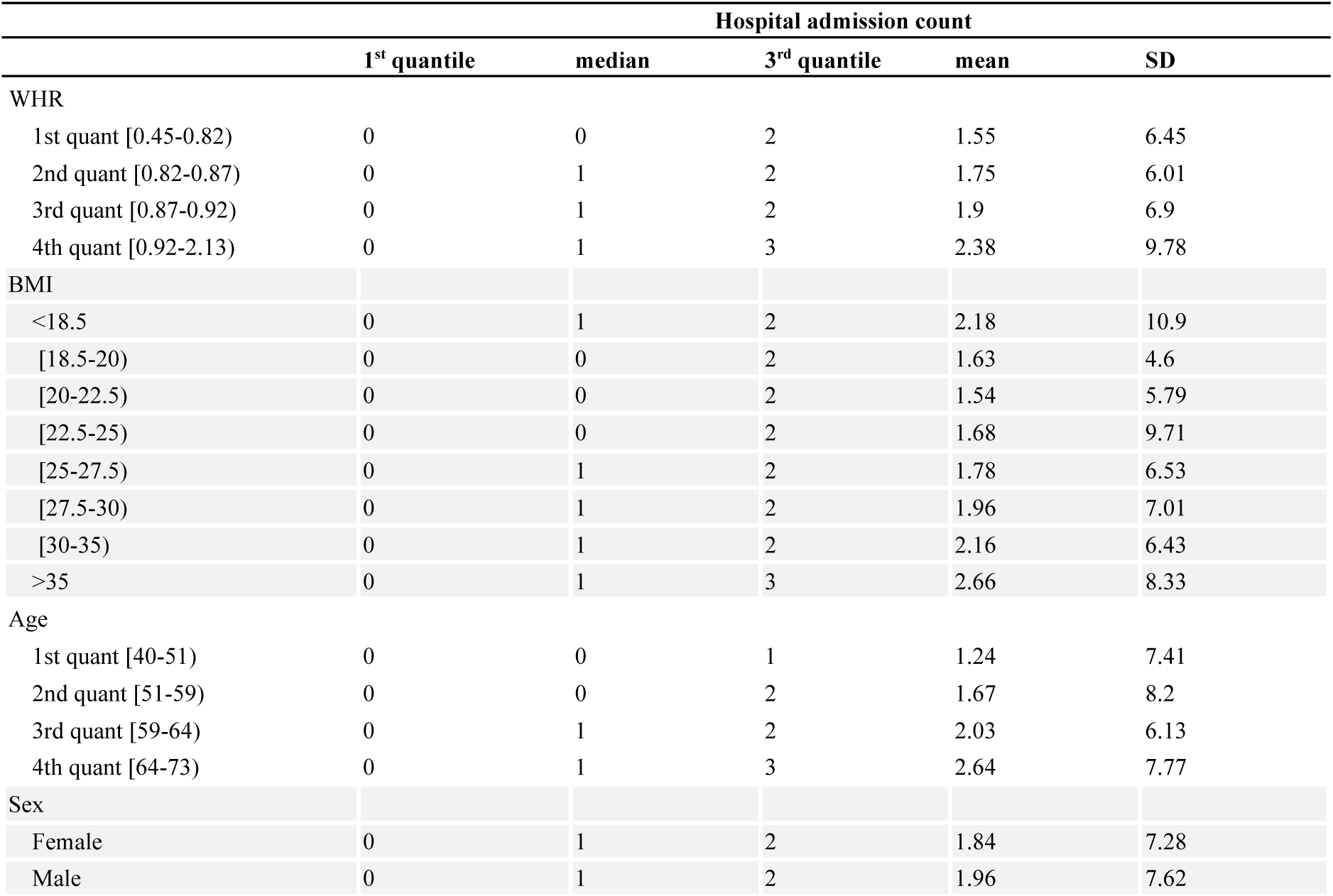
Hospital admission counts for 310471 patients per WHR quantiles, age quantiles, BMI categories and across gender. Shown are the 1^st^ quantile, median, 3^rd^ quantile, mean and SD.

|  | Hospital admission count |  |  |  |  |
| --- | --- | --- | --- | --- | --- |
|  | 1<sup>st</sup> quantile | median | 3<sup>rd</sup> quantile | mean | SD |
| WHR |  |  |  |  |  |
| 1st quant [0.45-0.82) | 0 | 0 | 2 | 1.55 | 6.45 |
| 2nd quant [0.82-0.87) | 0 | 1 | 2 | 1.75 | 6.01 |
| 3rd quant [0.87-0.92) | 0 | 1 | 2 | 1.9 | 6.9 |
| 4th quant [0.92-2.13) | 0 | 1 | 3 | 2.38 | 9.78 |
| BMI |  |  |  |  |  |
| <18.5 | 0 | 1 | 2 | 2.18 | 10.9 |
| [18.5-20) | 0 | 0 | 2 | 1.63 | 4.6 |
| [20-22.5) | 0 | 0 | 2 | 1.54 | 5.79 |
| [22.5-25) | 0 | 0 | 2 | 1.68 | 9.71 |
| [25-27.5) | 0 | 1 | 2 | 1.78 | 6.53 |
| [27.5-30) | 0 | 1 | 2 | 1.96 | 7.01 |
| [30-35) | 0 | 1 | 2 | 2.16 | 6.43 |
| >35 | 0 | 1 | 3 | 2.66 | 8.33 |
| Age |  |  |  |  |  |
| 1st quant [40-51) | 0 | 0 | 1 | 1.24 | 7.41 |
| 2nd quant [51-59) | 0 | 0 | 2 | 1.67 | 8.2 |
| 3rd quant [59-64) | 0 | 1 | 2 | 2.03 | 6.13 |
| 4th quant [64-73) | 0 | 1 | 3 | 2.64 | 7.77 |
| Sex |  |  |  |  |  |
| Female | 0 | 1 | 2 | 1.84 | 7.28 |
| Male | 0 | 1 | 2 | 1.96 | 7.62 |

### Observational multivariable analyses

Poisson multivariable regression models provided evidence that BMI and WHR were associated with an increase in yearly hospital admission rate (Table 3).

**Table 3.**
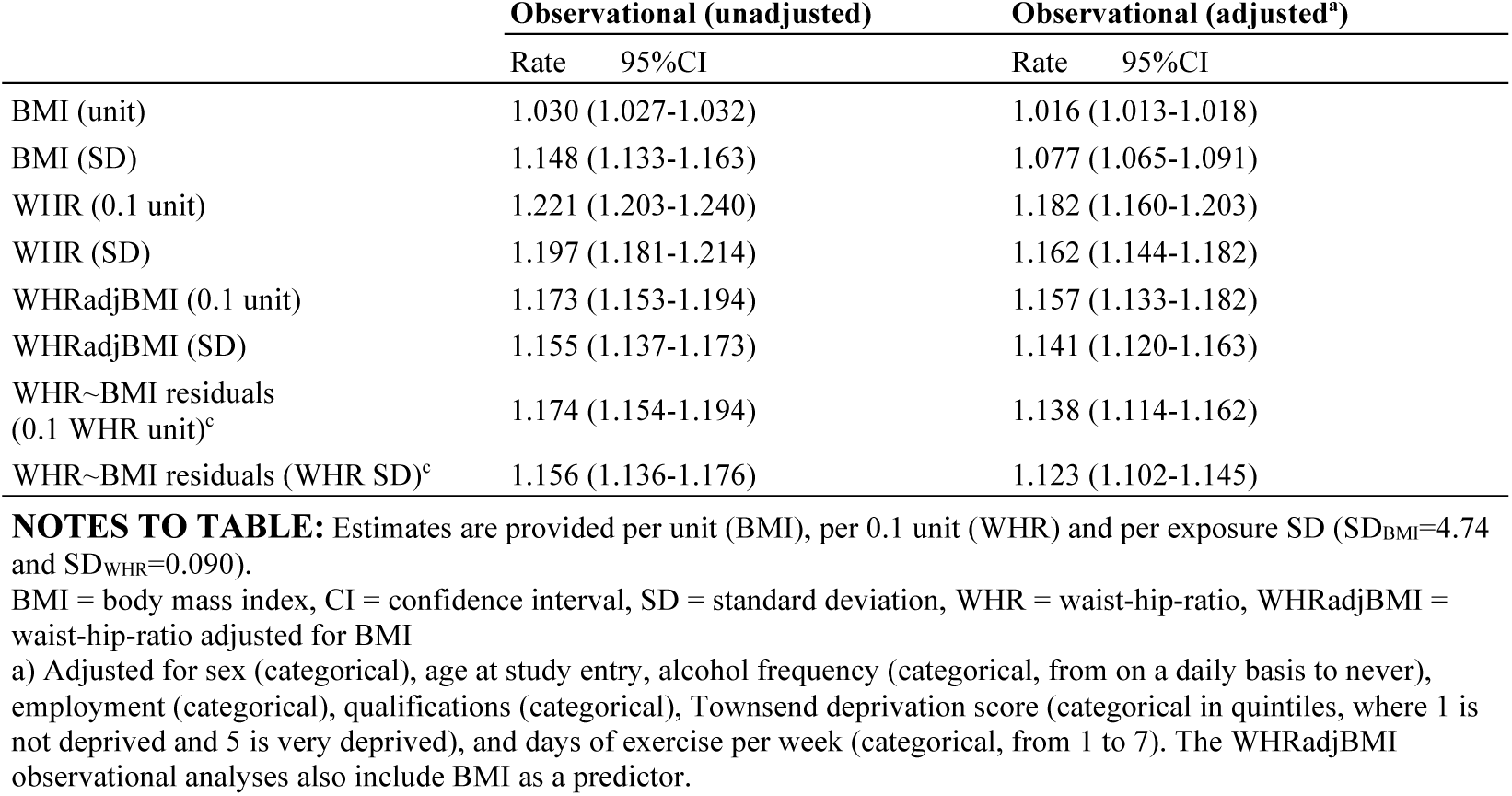
Observational multivariable analysis of the effect of BMI on yearly hospital admission rate per year in UK Biobank participants of White British ancestry (*N*=310,471).

Observational analyses regressing the outcome on both exposures simultaneously resulted in attenuated associations, with a 1.03-fold increase per BMI SD (95% CI: 1.02, 1.04) and a 1.14-fold increase (95% CI: 1.12, 1.16) per WHR SD, respectively (Table 5).

**Table 4.** Observational multivariable of the effect of BMI and WHR on yearly hospital admission rate

|  | Observational (unadjusted) |  | Observational (adjusted<sup>a</sup>) |  |
| --- | --- | --- | --- | --- |
|  | Rate | 95% CI | Rate | 95% CI |
| BMI (per unit) | 1.017 | (1.014-1.020) | 1.006 | (1.003-1.009) |
| BMI (per SD) | 1.084 | (1.068-1.100) | 1.029 | (1.015-1.043) |
| WHR (per 0.1 unit) | 1.173 | (1.153-1.194) | 1.158 | (1.134-1.182) |
| WHR (per SD) | 1.155 | (1.137-1.173) | 1.141 | (1.120-1.163) |

**NOTES TO TABLE:** Observational multivariable of the effect of BMI and WHR on yearly hospital admission rate in UK Biobank participants of White British ancestry ( $N=310471$ ). Rates and 95% confidence intervals (95% CI) are given. Estimates are provided per unit (BMI), per 0.1 unit (WHR) and per exposure SD ( $SD_{BMI}=4.74$ and $SD_{WHR}=0.090$ ).
BMI = body mass index, CI = confidence interval, SD = standard deviation, WHR = waist-hip-ratio
a) The observational analysis regresses the outcome directly on the exposures BMI and WHR simultaneously. The Poisson regression is adjusted for sex (categorical), age at study entry, alcohol frequency (categorical, from on a daily basis to never), employment (categorical), qualifications (categorical), Townsend deprivation score (categorical in quintiles, where 1 is not deprived and 5 is very deprived), and days of exercise per week (categorical, from 1 to 7);
Estimates (with corresponding 95% CIs) represent the fold increase in yearly hospital admission rate per BMI unit ( $1 \text{ kg/m}^2$ ) and SD ( $4.74 \text{ kg/m}^2$ ) and per 0.1 WHR unit and SD (0.090)

**Table 5.** Association between weighted GRS for BMI, WHR and WHRadjBMI with BMI and WHR in UK Biobank participants of White British ancestry (*N*=310471). Effect estimates are provided per unit (BMI), per 0.1 unit (WHR) and per SD (SD_BMI_=4.74 and SD_WHR_=0.090).

|  | Effect estimate (95% CI)<sup>a</sup> |  | P-value | R<sup>2</sup> % | F |
| --- | --- | --- | --- | --- | --- |
| *BMI GRS (76 SNPs)* |  |  |  |  |  |
| BMI (unit) | 0.112 | (0.109 – 0.115) | <5 x 10<sup>-324</sup> | 1.69 | 5326 |
| BMI (SD) | 0.0236 | (0.023 – 0.0243) | <5 x 10<sup>-324</sup> | 1.69 | 5326 |
| WHR (0.1 unit) | 0.0075 | (0.0069 – 0.0081) | <4.04 x 10<sup>-144</sup> | 0.210 | 654.1 |
| WHR (SD) | 0.0084 | (0.0077 – 0.009) | <4.04 x 10<sup>-144</sup> | 0.210 | 654.1 |
| *WHR GRS (39 SNPs)* |  |  |  |  |  |
| BMI (unit) | 0.0297 | (0.0256-0.0337) | <2.33 x 10<sup>-46</sup> | 0.066 | 204.4 |
| BMI (SD) | 0.0060 | (0.005-0.007) | <2.33 x 10<sup>-46</sup> | 0.066 | 204.4 |
| WHR (0.1 unit) | 0.0140 | (0.0132 – 0.0148) | <6.40 x 10<sup>-277</sup> | 0.406 | 1267 |
| WHR (SD) | 0.0156 | (0.0147 – 0.0164) | <6.40 x 10<sup>-277</sup> | 0.406 | 1267 |
| *WHRadjBMI GRS (48 SNPs)* |  |  |  |  |  |
| BMI (unit) | -0.024 | (-0.0278 – -0.0202) | <2.41 x 10<sup>-35</sup> | 0.050 | 154 |
| BMI (SD) | -0.005 | (-0.006 – -0.004) | <2.41 x 10<sup>-35</sup> | 0.050 | 154 |
| WHR (0.1 unit) | 0.0141 | (0.0134 – 0.0148) | <1.88 x 10<sup>-322</sup> | 0.474 | 1477 |
| WHR (SD) | 0.0157 | (0.0149 – 0.0165) | <1.88 x 10<sup>-322</sup> | 0.474 | 1477 |

**NOTES TO TABLE:** BMI = body mass index, CI = confidence interval, GRS = genetic risk score, SD = standard deviation, WHR = waist-hip-ratio, WHRadjBMI = waist-hip-ratio adjusted for BMI
a) Effect estimate, and corresponding P-value represent the change in BMI in units ( $\text{kg/m}^2$ ) and SD units ( $4.74 \text{ kg/m}^2$ ) and the change in WHR in 0.1 units and SD units (0.090) per BMI increasing allele (BMI GRS) and WHR increasing allele (WHR GRS, WHRadjBMI GRS)

For all exposures, estimates derived from adjusted models were lower than those from unadjusted models, but with overlapping confidence intervals.

### Association between GRS and exposures

Table 5 shows the associations between BMI and WHR and the BMI GRS, the WHR GRS, and the WHRadjBMI GRS, comprising 76, 39 and 48 SNPs, respectively. The first stage F statistics were large in all cases.

In UK Biobank participants of White British ethnicity, each unit increase in BMI GRS was associated with a 0.11kg/m^2^ higher BMI (95% CI: 0.11, 0.12), with the GRS explaining 1.69% of the variance. Each unit increase in WHR GRS was associated with a 0.01 higher WHR on the 0.10 unit scale (95% CI: 0.01, 0.01), explaining 0.41% of the variance, while each unit increase in WHRadjBMI GRS was associated with a 0.01 increase in WHR on the 0.10 unit scale (95% CI: 0.01, 0.02), explaining 0.47% of the variance. *F*-statistics indicated that each GRS was a strong instrument for Mendelian Randomization analyses (Table 5), with all *F*-statistics > 1,267.

### One-sample Mendelian Randomization analyses

One-sample Mendelian Randomization estimates of the effect of BMI, WHR and WHRadjBMI on hospital admission rates were obtained per exposure SD and unit increase. The IV regressions (Table 6), adjusted for age, sex and the first 40 genetic PCAs, yielded a 1.13-fold increase per BMI SD (95% CI: 1.02, 1.27) and a 1.26-fold increase per WHR SD (95% CI: 1.00, 1.58). Using the WHRadjBMI SNPs yielded a 1.22-fold increase per WHR SD (95% CI: 1.01, 1.47). Adjusting for BMI in the WHR regression, by using the residuals from a linear regression of WHR on BMI as an exposure, resulted in a reduced effect of 1.16 (95% CI: 0.97, 1.39) per SD. We include the adjusted observational estimates from Table 3 for comparison

**Table 6.** One-sample MR analyses of the effect of BMI (76 SNPs), WHR (39 SNPs) WHRadjBMI (48 SNPs) on yearly hospital admission rate per year in UK Biobank participants of White British ancestry (*N*=310471). Rates and 95% confidence intervals (95% CI) are given. Estimates are provided per unit (BMI), per 0.1 unit (WHR) and per exposure SD (SD_BM_I=4.74 and SD_WHR_=0.090).

|  | Observational (adjusted<sup>a</sup>) |  | IV (adjusted<sup>b</sup>) |  |
| --- | --- | --- | --- | --- |
|  | Rate | 95%CI | Rate | 95%CI |
| BMI (unit) | 1.016 | (1.013-1.018) | 1.027 | (1.003-1.051) |
| BMI (SD) | 1.077 | (1.065-1.091) | 1.134 | (1.015-1.267) |
| WHR (0.1 unit) | 1.182 | (1.160-1.203) | 1.287 | (0.997-1.661) |
| WHR (SD) | 1.162 | (1.144-1.182) | 1.255 | (0.997-1.580) |
| WHRadjBMI (0.1 unit) | 1.157 | (1.133-1.182) | 1.242 | (1.010-1.529) |
| WHRadjBMI (SD) | 1.141 | (1.120-1.163) | 1.216 | (1.009-1.466) |
| WHR~BMI residuals (0.1 WHR unit)<sup>c</sup> | 1.138 | (1.114-1.162) | 1.180 | (0.965-1.442) |
| WHR~BMI residuals (WHR SD)<sup>c</sup> | 1.123 | (1.102-1.145) | 1.161 | (0.968-1.391) |

BMI = body mass index, CI = confidence interval, IV = instrumental variable, SD = standard deviation, WHR = waist-hip-ratio, WHRadjBMI = waist-hip-ratio adjusted for BMI
a) Adjusted for sex (categorical), age at study entry, alcohol frequency (categorical, from on a daily basis to never), employment (categorical), qualifications (categorical), Townsend deprivation score (categorical in quintiles, where 1 is not deprived and 5 is very deprived), and days of exercise per week (categorical, from 1 to 7). The WHRadjBMI observational analyses also include BMI as a predictor.
b) Adjusted for sex, age at study entry, and 40 PCAs
c) Residuals from linear WHR on BMI regressions are used as an exposure with the WHRadjBMI SNPs as instruments

The multivariable one-sample Mendelian Randomization analysis (Table 7) showed no strong evidence for an independent effect of BMI on hospital admissions, with a fold increase of 1.04 (95% CI: 0.93, 1.15) per BMI SD, when controlling for WHR. Again, we include the adjusted observational effect estimates (from Table 4) for comparison.

**Table 7.** Observational multivariable and one-sample multivariable MR analyses of the effect of BMI and WHR on yearly hospital admission rate in UK Biobank participants of White British ancestry (*N*=310471). Rates and 95% confidence intervals (95% CI) are given. Estimates are provided per unit (BMI), per 0.1 unit (WHR) and per exposure SD (SD_BMI_=4.74 and SD_WHR_=0.090).

|  | Observational (adjusted<sup>a</sup>) |  | IV (adjusted<sup>b</sup>) |  |
| --- | --- | --- | --- | --- |
|  | Rate<sup>c</sup> | 95%CI | Rate<sup>c</sup> | 95%CI |
| BMI (per unit) | 1.006 | (1.003-1.009) | 1.007 | (0.985-1.031) |
| BMI (per SD) | 1.029 | (1.015-1.043) | 1.035 | (0.930-1.153) |
| WHR (per 0.1 unit) | 1.158 | (1.134-1.182) | 1.354 | (1.041-1.761) |
| WHR (per SD) | 1.141 | (1.120-1.163) | 1.314 | (1.037-1.665) |

BMI = body mass index, CI = confidence interval, GRS = genetic risk score, SD = standard deviation, WHR = waist-hip-ratio
a) The observational analysis regresses the outcome directly on the exposures BMI and WHR simultaneously. The Poisson regression is adjusted for sex (categorical), age at study entry, alcohol frequency (categorical, from on a daily basis to never), employment (categorical), qualifications (categorical), Townsend deprivation score (categorical in quintiles, where 1 is not deprived and 5 is very deprived), and days of exercise per week (categorical, from 1 to 7);
b) Adjusted for sex, age at study entry, and 40 PCAs
c) Estimates (with corresponding 95% CIs) represent the fold increase in yearly hospital admission rate per BMI unit ( $1 \text{ kg/m}^2$ ) and SD ( $4.74 \text{ kg/m}^2$ ) and per 0.1 WHR unit and SD (0.090)

Conversely, there was evidence for an independent effect of WHR on hospital admissions, with a fold increase of 1.31 (95% CI: 1.04, 1.67) per WHR SD. Instrument strength, as assessed using the conditional Sanderson-Windmeijer *F*-statistic (Sanderson et al., 2019), was sufficient for the multivariable Mendelian Randomization analysis, with *F*-statistics for BMI of 38.50 and for WHR of 22.89.

### Two-sample Mendelian Randomization analyses

The IVW estimator showed evidence for a causal effect of all three exposures on hospital admissions (Table 8), at magnitudes consistent with the just-identified one-sample Mendelian Randomization results.

**Table 8.** Two-sample MR analysis of hospital admission rate per year in UK Biobank

|  |  | IVW (random effects, exact weights) | MR-Egger |  | Penalized weighted median | Weighted mode |
| --- | --- | --- | --- | --- | --- | --- |
|  |  |  | Intercept | Slope |  |  |
| BMI (unit) | Rate<sup>a</sup><br>(95% CI) | 1.020<br>(1.002-1.038) | 1.001<br>(0.999-1.002) | 0.994<br>(0.943-1.048) | 1.020<br>(0.986-1.055) | 1.019<br>(0.981-1.059) |
| BMI (SD) | Rate<sup>a</sup><br>(95% CI) | 1.098<br>(1.009-1.194) | 1.004<br>(0.997-1.011) | 0.973<br>(0.759-1.247) | 1.095<br>(0.930-1.290) | 1.095<br>(0.915-1.312) |
| WHR (0.1 unit) | Rate<sup>a</sup><br>(95% CI) | 1.223<br>(1.062-1.407) | 1.000<br>(0.987-1.014) | 1.208<br>(0.648-2.253) | 1.265<br>(0.982-1.630) | 1.229<br>(0.864-1.748) |
| WHR (SD) | Rate<sup>a</sup><br>(95% CI) | 1.199<br>(1.054-1.364) | 1.000<br>(0.987-1.014) | 1.185<br>(0.676-2.079) | 1.236<br>(0.985-1.550) | 1.204<br>(0.867-1.167) |
| WHRadjBMI (0.1 unit) | Rate<sup>a</sup><br>(95% CI) | 1.168<br>(1.030-1.326) | 1.008<br>(0.995-1.021) | 0.857<br>(0.506-1.453) | 1.138<br>(0.933-1.389) | 1.084<br>(0.798-1.475) |
| WHRadjBMI (SD) | Rate<sup>a</sup><br>(95% CI) | 1.151<br>(1.028-1.287) | 1.008<br>(0.995-1.021) | 0.870<br>(0.541-1.400) | 1.124<br>(0.941-1.343) | 1.076<br>(0.819-1.414) |

**NOTES TO TABLE:** MR-Egger (random effects), IVW (random effects, exact weights), weighted median and weighted mode analyses of BMI (64 SNPs), WHR (34 SNPs) and WHRadjBMI (45 SNPs) on hospital admission rate per year in UK Biobank participants of White British ancestry. SNPs with an LD $R^2 < 0.001$ have been retained. Rates are given per exposure unit and exposure SD ( $SD_{BMI}=4.6$ , $SD_{WHR}=0.07$ ) and 95% confidence intervals (95% CI) are provided.
BMI = body mass index, CI = confidence interval, IVW = inverse variance weighted, MR = Mendelian randomization, SD = standard deviation, WHR = waist-hip-ratio, WHRadjBMI = waist-hip-ratio adjusted for BMI
a) Adjusted for sex, age and the first 40 genetic principle components. Estimates (with corresponding 95% CIs) represent the fold increase in yearly hospital admission rate per BMI unit (1 kg/m<sup>2</sup>) and SD (4.6 kg/m<sup>2</sup>) and per 0.1 WHR unit and SD (0.07)

For BMI, we observed a fold increase of 1.10 (95% CI: 1.01, 1.19) per SD, for WHR a fold increase of 1.20 (95% CI: 1.05, 1.36) per SD and for WHRadjBMI a fold increase of 1.15 (95% CI: 1.03, 1.29) per SD. The penalized weighted median and weighted mode yielded near identical point estimates, albeit with wider confidence intervals that included the null.

The widest confidence intervals were observed for the Mendelian Randomization-Egger estimates, as is usually the case given the lower power of this estimator since it estimates twice the number of parameters (both the intercept and the slope coefficient) compared to the other estimates. For WHR, the Mendelian Randomization-Egger point estimate was comparable to the previous estimates, it was markedly lower for BMI and WHRadjBMI. The Mendelian Randomization-Egger intercept indicated no directional pleiotropy.

Heterogeneity was assessed using Rücker’s Q (Q_R_) and Cochran’s Q (Q_C_) For all three exposures, the Q-statistics were smaller than the number of SNPs used for estimation (Q_R,BMI_=51.29, Q_C,BMI_=52.06, Q_R,WHR_=28.64, Q_C,WHR_=28.92, Q_R,WHRadj_=43.80, Q_C,WHRadj_=44.01). Despite the lack of substantial heterogeneity, we performed a sensitivity analysis to investigate the potential presence of pleiotropy driven by outlying SNPs. Individual SNPs were identified in a visual inspection of leave-one-out plots (Figures S1-S3) and removed, with 3, 4 and 4 SNPs excluded for BMI, WHR, and WHRadjBMI, respectively. The two-sample MR analyses were repeated using the reduced set of SNPs, yielding comparable point estimates and confidence intervals overlapping with those from the original analyses (Table S2).

A multivariable two-sample IVW analysis of BMI and WHR simultaneously (Table 10) yielded no strong evidence for an association of BMI and the yearly hospital admission rate, with a fold increase of 0.99 per BMI SD (95% CI: 0.85, 1.14), but indicated a strong association between WHR and hospital admission rate, with a fold increase of 1.30 per WHR SD (95% CI: 1.02, 1.65).

**Table 10.** Multivariable two-sample MR IVW estimates for the effect of BMI and WHR (70 SNPs) on yearly hospital admission rate in UK Biobank participants of White British ancestry.

|  | Rate | 95% CI |
| --- | --- | --- |
| BMI (unit) | 0.997 | (0.966-1.029) |
| BMI (SD) | 0.986 | (0.850-1.143) |
| WHR (0.1 unit) | 1.335 | (1.024-1.739) |
| WHR (SD) | 1.297 | (1.022-1.647) |

**NOTES TO TABLE:** SNPs with an LD $R^2 < 0.001$ were retained. Rates are given per exposure unit and exposure SD ( $SD_{BMI}=4.6$ , $SD_{WHR}=0.07$ ) and 95% confidence intervals (95% CI) are provided.
BMI = body mass index, CI = confidence interval, IVW = inverse variance weighted, LD = linkage disequilibrium, MR = Mendelian randomization, SD = standard deviation, WHR = waist-hip-ratio
a) Adjusted for sex, age and the first 40 genetic principal components. Estimates (with corresponding 95% CIs) represent the fold increase in yearly hospital admission rate per BMI unit (1 kg/m<sup>2</sup>) and SD (4.6 kg/m<sup>2</sup>) and per 0.1 WHR unit and SD (0.07)

The gene-exposure associations and gene-outcome associations for the univariable two-sample Mendelian Randomization analyses are given in Supplementary Tables S10, S11 and S12 for BMI, WHR, and WHRadjBMI, respectively. The same quantities are provided for the multivariable two-sample analysis in Supplementary Table S13.

### Comparison across methods

Figure 2 summarizes the effect of each exposure on yearly hospital admission rate, as estimated from traditional multivariable analyses, and one- and two-sample Mendelian Randomization analyses.

**Figure 2.**
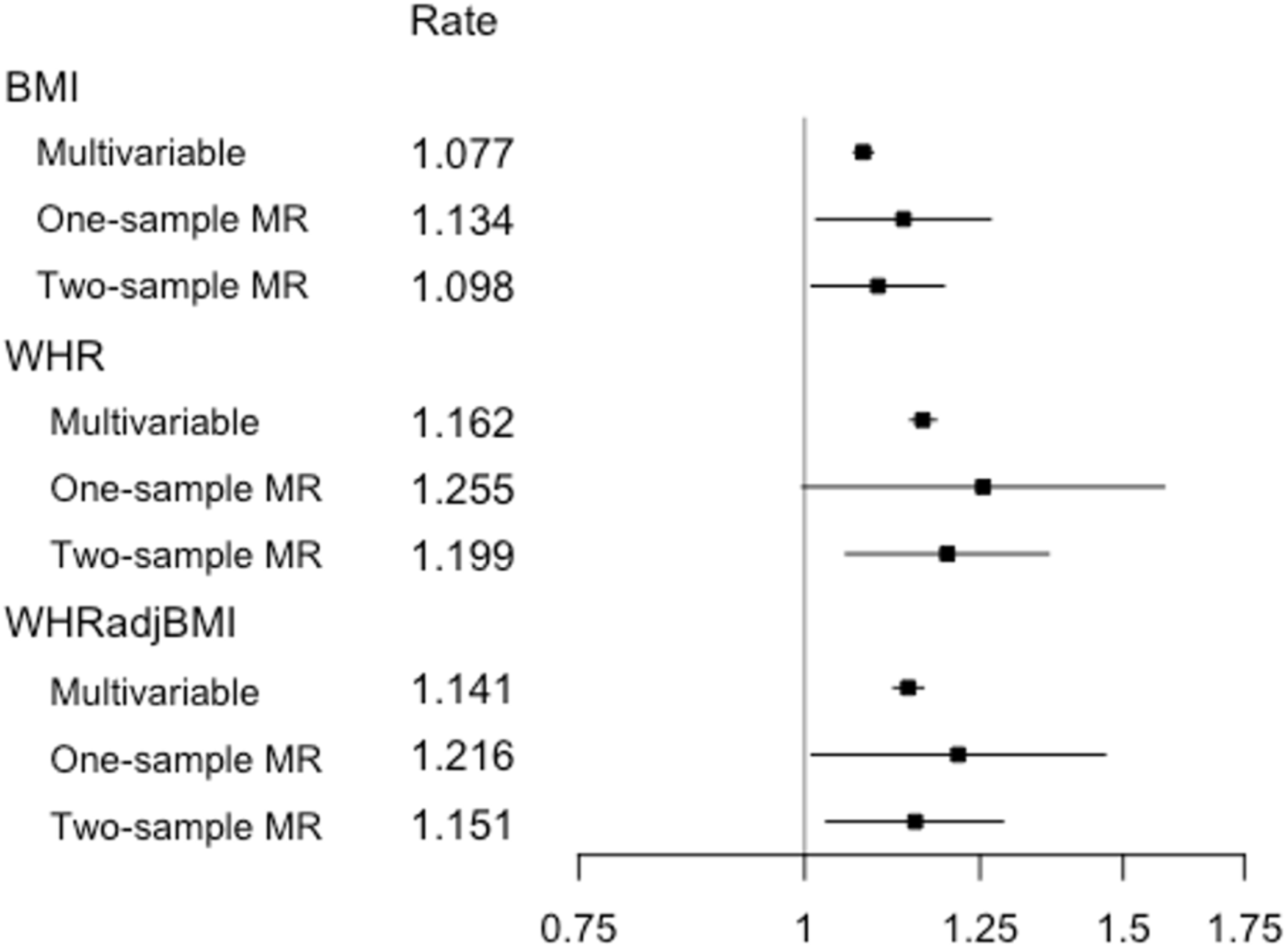
Estimates from multivariable observational analyses, one-sample Mendelian Randomization analyses and two-sample Mendelian Randomization IVW analyses for exposures BMI, WHR and WHRadjBMI per SD unit. **NOTES TO FIGURE**: Shown are point estimates alongside 95% CIs for the effect of exposure on yearly hospital admission rate. The results are plotted on the log scale, to ensure symmetrical CIs and comparability of magnitude across estimates. Rate estimates and x-axis values are given on the rate scale. All Mendelian Randomization analyses were adjusted for age, sex and the first 40 genetic PCAs. The multivariable observational analyses were adjusted for a range of baseline patient characteristics (Table S2).

For all exposures, we observed a higher point estimate for the one-sample Mendelian Randomization analyses when compared to the traditional analyses. On the whole, the estimated effects were consistent across estimators, with overlapping confidence intervals, and comparatively wider confidence intervals for WHR and WHRadjBMI.

A similar pattern is observed in Figure 3, which summarizes the multivariable estimator results. Once more, the confidence intervals for both exposures overlapped, but with the BMI point estimates grouped more closely. In contrast to the univariable analyses summarized in Figure 2, the one- and two-sample BMI estimate confidence intervals now included the null.

**Figure 3.**
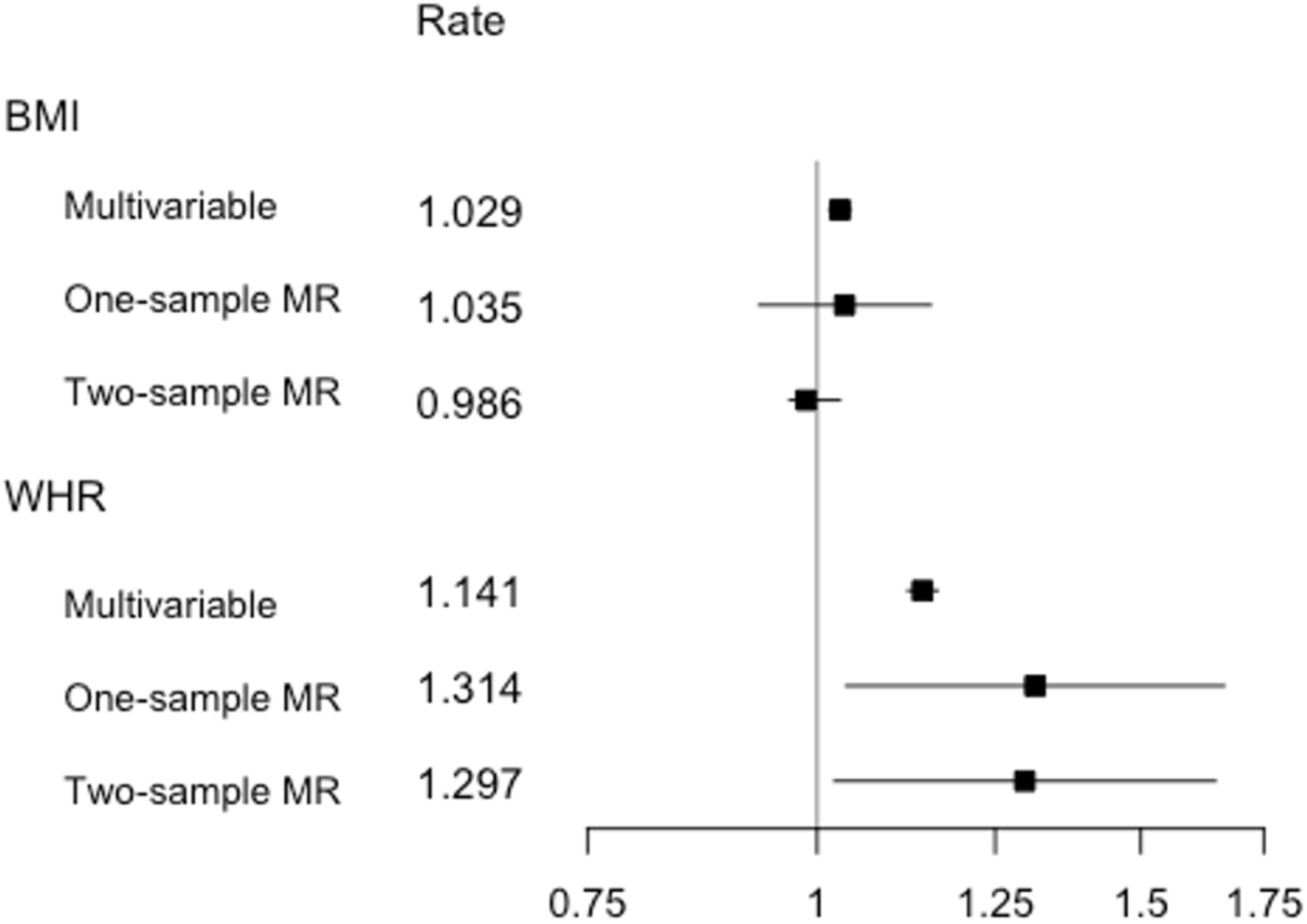
Estimates from multivariable observational analyses, multivariable one-sample Mendelian Randomization analyses and multivariable two-sample Mendelian Randomization IVW analyses for exposures BMI and WHR per SD unit. **NOTES TO FIGURE:** Shown are point estimates alongside 95% CIs for the effect of exposure on yearly hospital admission rate. The results are plotted on the log scale, to ensure symmetrical CIs and comparability of magnitude across estimates. Rate estimates and x-axis values are given on the rate scale. Mendelian Randomization analyses were adjusted for age, sex and the first 40 genetic PCAs. The multivariable observational analyses were adjusted for a range of baseline patient characteristics (Table S2), with the BMI estimate adjusted for WHR and vice versa.

## Discussion

To our knowledge, this is the first time all-cause hospital admissions have been modeled using Mendelian Randomization methods, a methodology less affect by omitted variable bias and reverse causation than other methods SEARCH (G. Davey Smith & Hemani, 2014; Haycock et al., 2016). Our Mendelian Randomization results, based on analysis of more than 550,000 hospital admissions measured in over 300,000 UK Biobank participants, supported the presence of causal effect of a higher BMI, WHR and WHRadjBMI on an increased hospital admission risk using one- and two-sample Mendelian Randomization methods, in both a univariable and multivariable Mendelian Randomization framework.

In the multivariable one-sample and two-sample Mendelian Randomization analysis, a relatively stronger positive effect was observed for WHR than for BMI, suggesting that the relationship between adiposity and hospital admissions may be driven by a detrimental distribution of fat and adipose tissue rather than by BMI itself. WHR has been little investigated previously in the context of hospital admissions, and our results emphasize the relevance of considering WHR as a measure of adiposity in addition to BMI in this context. Results from sensitivity analyses that relaxed the exclusion restriction were broadly concordant across methods relying on different assumptions, suggesting that the same causal effect was being identified.

A positive association of BMI and all-cause hospital admissions has previously been shown in observational studies investigating populations from the UK (Kent, Green, et al., 2017; O’Halloran, 2020), Australia (Korda et al., 2015), Canada (Chen et al., 2007), Italy (Migliore et al., 2013), and the USA (Buys et al., 2014; Han et al., 2009). An observational study of approximately 1.09 million UK women found a yearly hospital admission rate increase of 1.12 (95% CI: 1.12, 1.13) for every 5kg/m^2^ increase in BMI (Reeves et al., 2014). In just over 300,000 individuals, we observed estimates amounting to rate coefficients of 1.02^5^ = 1.08 (95% CI: 1.07, 1.10), 1.14 (95% CI: 1.02, 1.28) and 1.10 (95% CI: 1.01, 1.21) from the multivariable observational, one-sample Mendelian Randomization and two-sample Mendelian Randomization analyses, respectively, giving effect estimates of a magnitude comparable to those observed previously in UK women from observational studies.

Another observational study, examining the association between BMI and hospital admissions in 451,320 UK Biobank participants, found increases in yearly hospital admission rates, measured per 2kg/m^2^ BMI, of 1.06 (95% CI: 1.05, 1.07) and 1.06 (95% CI: 1.05, 1.07) for male and female never-smokers, respectively (O’Halloran, 2020). We observed comparable estimates, with increases across both genders, per 2kg/m^2^ BMI, of 1.02^2^=1.03 (95% CI: 1.03, 1.04), 1.05 (95% CI: 1.01, 1.11), and 1.04 (95% CI: 1.00, 1.08), for the multivariable observational, one-sample Mendelian Randomization and two-sample Mendelian Randomization analyses, respectively.

While the effect of BMI/WHR on hospital admission rate has not previously been studied in this context, the effect of BMI on hospital cost was examined in a two-sample Mendelian Randomization analysis, using data from UK Biobank, which largely overlaps with our own study population (P. Dixon et al., 2020). A positive causal effect of BMI on hospital cost was found in that study, in line with our own observation of increased hospital admissions for higher BMI.

### Limitations

Mendelian Randomization methods make it possible to avoid certain biases common to traditional epidemiological studies, but also face limitations, both in terms of interpretation and in terms of potential alternative sources of bias. When interpreting the results, it should be noted that Mendelian Randomization does not estimate an average treatment effect, but rather a local average treatment effect (LATE) instead, under the assumption that the effect of IV on treatment for all IVs is in the same direction for all subjects – the condition of monotonicity (Von Hinke et al., 2016). In context of this particular study, this means that we estimate the effect of WHR and BMI in those subjects whose WHR/BMI exposure values differ on varying the levels of the respective IVs, under the condition that the change occurs in the same direction for all participants. This is probably a reasonable assumption.

As the IVs are comprised of genetic markers, which are ‘assigned’ at conception, the estimated LATE is a measure of the effects of a lifelong exposure to BMI-increasing alleles/WHR-increasing alleles. Additionally, we should note that, for all three exposures, a relatively modest percentage of variance is explained by the genetic variants, reducing statistical power for detecting the effect of a change in BMI/WHR and consequently less precise (Haycock et al., 2016) (albeit potentially less biased) estimates.

A key interpretive assumption of Mendelian Randomization is that of gene-environment equivalence: that genetically influenced BMI and WHR will have the same effect on hospital admission risk as, for example, adiposity modified by diet and/or exercise. The included SNPs, however, may not meet the stable unit treatment assumption (SUTVA) (S. Burgess & Thompson, 2015) and as such, our estimates of the effect of BMI and WHR will not necessarily be representative of the increase or reduction in hospital admission rate when adiposity is altered through interventions.

We were limited to inpatient hospital admissions as the source of hospital data linked to UK Biobank at the time of writing. It is possible that other forms of hospital and primary care may substitute for inpatient care, in which case our estimates may overstate the effect of adiposity on overall healthcare admissions and care episodes. On the other hand, if inpatient care is complementary to other forms of care, then we may have understated effect sizes. In practice, both influences may be present. Our results are best interpreted in relation to our outcome of inpatient admissions.

While UK Biobank is a unique and high-quality source, the underlying demographic structure of the data imposes various restrictions in terms of generalizability. The UK Biobank sample is healthier and wealthier than the population from which it is drawn and consequently is likely to not be representative of the wider UK. Indeed, we observed lower rates of mortality than in the general population (Bycroft et al., 2018), better health-related behaviour and a higher level of education (Fry et al., 2017).

As less healthy individuals are less likely to participate in the study, the observed sample may be subject to selection bias (Glymour, 2006; Munafò, Tilling, Taylor, Evans, & Davey Smith, 2018; Spirtes, Glymour, & Scheines, 1993), which will impact the estimates obtained from both conventional multivariable analyses and Mendelian Randomization analyses – where estimates obtained may even be an underestimation of the effect of adiposity on hospital admissions in the general population. It should also be noted that current analyses have been limited to individuals of White British ancestry, and as such the results will not necessarily generalize to other ancestral groups.

A potential further limitation is the possibility of cohort effects. UK Biobank participants are aged 39-72 years, giving rise to range of birth cohorts in our data sample. There is some evidence from other sources to suggest that SNPs exert a greater influence on BMI for those born in more recent decades, possibly because of an increasingly obesogenic environment (Hartwig, Davies, & Davey Smith, 2018; Walter, Mejía-Guevara, Estrada, Liu, & Glymour, 2016). In this case, our estimates may under-estimate the impact of adiposity on hospital admissions, other things being equal.

Additional sources of bias that should be considered in the context of Mendelian Randomization and the UK Biobank data are assortative mating (Hartwig et al., 2018; Jacobson, Torgerson, Sjöström, & Bouchard, 2007; Morris, Davies, Hemani, & Davey Smith, 2019; Tenesa, Rawlik, Navarro, & Canela-Xandri, 2016), confounding due to population stratification bias (Brumpton et al., 2020; Haworth et al., 2019; Koellinger & de Vlaming, 2019; Morris et al., 2019) and the presence of dynastic effects (Brumpton et al., 2020; Fletcher, 2011; Morris et al., 2019), which have all been implicated with respect to BMI previously. While we attempted to minimize the impact of population stratification with the inclusion of 40 genetic principle components, latent data structure may still exist for some exposures such as BMI (Haworth et al., 2019). Within-family analysis may address some of these issues (Brumpton et al., 2020; Davies et al., 2019), although statistical power for this form of modelling is limited given relatively modest available sample sizes of related individuals in UK Biobank (Howe et al., 2020).

## Conclusion

This study describes the first Mendelian Randomization analysis to estimate the causal effect of BMI, WHR and WHRadjBMI on yearly hospital admission rates. Results supported the causal role of greater adiposity in increasing the risk of hospital admissions. Causal point estimates were larger than those obtained from conventional observational models, further emphasizing the necessity of exploring policies intended to address adverse adiposity profiles.

Multivariable Mendelian Randomization analyses suggested that the effect of BMI on hospital admission rates may be mediated by WHR and that an unfavourable fat distribution may drive the relationship between increased adiposity and higher hospital admission rates. Additionally, we demonstrate that a non-standard outcome like hospital admission counts can be successfully modeled using Mendelian Randomization methods, both in a one-sample and two-sample framework, by replacing the second stage regression (modeling the gene-outcome association) with a Poisson regression.

## Data Availability

All data used is available from UK biobank. The outcome of interest, hospital admission count, was derived using electronic health records linked to the UK Biobank study. Code to define the admissions variable is available from https://github.com/pdixon-econ/admissions-biobank.

## Acknowledgments

This research has been conducted using the UK Biobank Resource as part of application numbers 16391 and 29294.

