## Supplement for "Mendelian Randomization analysis of the causal impact of body mass index and waist-hip ratio on rates of hospital admission"

### Appendix

#### Supplementary tables and figures

*Tables*

**Table S1.** Cross-table of participant counts across eight exclusion criteria. 1
**Table S2.** Two-sample MR analyses; MR-Egger, IVW, weighted median and weighted mode analyses of BMI, WHR and WHRadjBMI on yearly hospital admission rate: a sensitivity analysis on exclusion of potentially pleiotropic SNPs 2
**Table S3.** BMI-SNP and hospital admission count-SNP associations 3
**Table S4.** WHR-SNP and hospital admission count-SNP associations 5
**Table S5.** WHRadjBMI-SNP and hospital admission count-SNP associations 7
**Table S6.** BMI-SNP, WHR-SNP and hospital admission count-SNP associations as used for the multivariable two-sample MR analysis. 9

*Figures*

**Figure S1.** Plots for two-sample MR analysis of BMI effect on yearly hospital admission rate: A) Cochran's Q against Rücker's Q; B) SNP effects on log scale 11
**Figure S2.** Plots for two-sample MR analysis of WHR effect on yearly hospital admission rate: A) Cochran's Q against Rücker's Q; B) SNP effects on log scale 12
**Figure S3.** Plots for two-sample MR analysis of WHRadjBMI effect on yearly hospital admission rate: A) Cochran's Q against Rücker's Q; B) SNP effects on log scale 13

**Table S1.** Cross-table of participant count across eight exclusion criteria. Total participant count per exclusion criteria is given on the diagonal, with the remaining table entries giving the number of participants meeting any two such criteria (e.g. 345 participants failed the standard inclusion criteria and were of non-white British ancestry). A total of 154901 participants match one or more exclusion criteria and are considered ineligible for analysis

| *Exclusion criterion* | Incorrect admission information[a] | Failed standard inclusion criteria[b] | Non-white British ancestry | Highly related[c] | Minimally related[d] | Not genotyped[e] | No BMI/WHR measurements | No PCA information |
|---|---|---|---|---|---|---|---|---|
| Incorrect admission information[a] | **34** | 0 | 1 | 0 | 1 | 27 | 1 | 27 |
| Failed standard inclusion criteria[b] | | **1675** | 345 | 0 | 100 | 899 | 8 | 0 |
| Non-white British ancestry | | | **73278** | 7 | 7431 | 225 | 415 | 0 |
| Highly Related[c] | | | | **8** | 0 | 0 | 0 | 0 |
| Minimally related[d] | | | | | **73893** | 0 | 255 | 0 |
| Not Genotyped[e] | | | | | | **13797** | 796 | 12898 |
| No BMI/WHR measurements | | | | | | | **2454** | 794 |
| No PCA information | | | | | | | | 12899 |

a) admissions prior to study start date, post death/censoring date or registered death prior to study start; b) individuals that have a mismatch between genetically inferred and reported gender, individuals with sex chromosome types putatively different from XX or XY and individuals that are outliers in heterozygosity and missing rate; c) individuals related to more than 200 other participants; d) on exclusion a maximal set of unrelated individuals is retained; ) not genotyped for the exposures of interest (BMI, WHR, WHRadjBMI)

**Table S2.** MR-Egger (random effects), IVW (random effects, exact weights), weighted median and weighted mode analyses of BMI (64 SNPs), WHR (34 SNPs) and WHRadjBMI (45 SNPs) on hospital admission rate per year in UK Biobank participants of White British ancestry. SNPs with an LD $R^2 < 0.001$ have been retained. Rates are given per exposure unit and exposure SD ($SD_{BMI}$=4.6, $SD_{WHR}$=0.07) and 95% confidence intervals (95% CI) are provided. Outliers identified in Figures S1, S2 and S3 were excluded as a sensitivity analysis (3, 4 and 4 SNPs for BMI, WHR and WHRadjBMI, respectively).

| | | IVW (random effects, exact weights) | MR-Egger | | Penalized weighted median | Weighted mode |
|---|---|---|---|---|---|---|
| | | | Intercept | Slope | | |
| BMI (unit) | Rate[a] (95% CI) | 1.018 (1.002-1.035) | 1.004 (0.997-1.011) | 0.992 (0.941-1.046) | 1.019 (0.983-1.058) | 1.020 (0.978-1.063) |
| BMI (SD) | Rate[a] (95% CI) | 1.089 (1.011-1.174) | 1.004 (0.997-1.011) | 0.963 (0.750-1.236) | 1.098 (0.931-1.294) | 1.098 (0.905-1.329) |
| WHR (0.1 unit) | Rate[a] (95% CI) | 1.166 (1.031-1.320) | 1.000 (0.986-1.017) | 1.115 (0.550-2.258) | 1.251 (0.978-1.600) | 1.234 (0.855-1.782) |
| WHR (SD) | Rate[a] (95% CI) | 1.149 (1.029-1.283) | 1.000 (0.986-1.017) | 1.103 (0.584-2.084) | 1.224 (0.975-1.536) | 1.209 (0.861-1.698) |
| WHRadjBMI (0.1 unit) | Rate[a] (95% CI) | 1.137 (1.024-1.264) | 1.002 (0.988-1.015) | 1.070 (0.624-1.834) | 1.139 (0.938-1.384) | 1.089 (0.792-1.497) |
| WHRadjBMI (SD) | Rate[a] (95% CI) | 1.123 (1.015-1.243) | 1.002 (0.988-1.015) | 1.063 (0.654-1.728) | 1.125 (0.945-1.338) | 1.080 (0.792-1.473) |

BMI = body mass index, CI = confidence interval, IVW = inverse variance weighted, LD = linkage disequilibrium, MR = Mendelian randomization, SD = standard deviation, WHR = waist-hip-ratio

a) Adjusted for sex, age and the first 40 genetic principle components. Estimates (with corresponding 95% CIs) represent the fold increase in yearly hospital admission rate per BMI unit (1 $kg/m^2$) and SD (4.6 $kg/m^2$) and per 0.1 WHR unit and SD (0.07)

**Table S3.** BMI-SNP and hospital admission count-SNP associations as used for the univariate two-sample MR analysis of the effect of BMI on yearly hospital admission rate. The BMI-SNP association coefficients were used as external weights for the genetic risk score in the one-sample MR analysis.

| | | | | **BMI-SNP associations (European ancestry GIANT consortium meta-analysis, Locke *et al.* (2016)** | | | | **Hospital admission count – SNP associations (UK Biobank)** | | | |
|---|---|---|---|---|---|---|---|---|---|---|---|
| **SNP** | **EA[b]** | **OA** | **EAF** | **β[c]** | **SE** | ***N*** | ***P*** | **β[d]** | **SE** | ***N*** | ***P*** |
| rs1000940 | G | A | 0.320 | 0.019 | 0.003 | 321836 | 1.28E-08 | -0.012 | 0.011 | 310537 | 9.25E-09 |
| rs10132280 | C | A | 0.682 | 0.023 | 0.003 | 321797 | 1.14E-11 | 0.003 | 0.011 | 310537 | 1.76E-01 |
| rs1016287 | T | C | 0.287 | 0.023 | 0.003 | 321969 | 2.25E-11 | 0.015 | 0.011 | 310537 | 9.98E-14 |
| rs10182181 | G | A | 0.462 | 0.031 | 0.003 | 321759 | 8.78E-24 | -0.009 | 0.010 | 310537 | 1.11E-06 |
| rs10733682 | A | G | 0.478 | 0.017 | 0.003 | 320727 | 1.83E-08 | -0.003 | 0.010 | 310537 | 1.24E-01 |
| rs10938397 | G | A | 0.434 | 0.040 | 0.003 | 320955 | 3.21E-38 | -0.004 | 0.010 | 310537 | 5.44E-02 |
| rs10968576 | G | A | 0.320 | 0.025 | 0.003 | 322061 | 6.61E-14 | -0.001 | 0.011 | 310537 | 6.46E-01 |
| rs11030104 | A | G | 0.792 | 0.041 | 0.004 | 322103 | 5.56E-28 | 0.014 | 0.013 | 310537 | 1.43E-09 |
| rs11057405 | G | A | 0.901 | 0.031 | 0.006 | 314111 | 2.02E-08 | 0.009 | 0.017 | 310537 | 3.95E-03 |
| rs11126666[a] | A | G | 0.283 | 0.021 | 0.003 | 321979 | 1.33E-09 | -0.007 | 0.012 | 310537 | 1.11E-03 |
| rs11165643 | T | C | 0.583 | 0.022 | 0.003 | 320730 | 2.07E-12 | 0.012 | 0.011 | 310537 | 5.15E-10 |
| rs11191560[a] | C | T | 0.089 | 0.031 | 0.005 | 321893 | 8.45E-09 | 0.020 | 0.019 | 310537 | 1.45E-08 |
| rs11583200[a] | C | T | 0.396 | 0.018 | 0.003 | 322095 | 1.48E-08 | -0.012 | 0.011 | 310537 | 1.10E-10 |
| rs1167827 | G | A | 0.553 | 0.020 | 0.003 | 306238 | 6.33E-10 | -0.004 | 0.010 | 310537 | 4.97E-02 |
| rs11688816[a] | G | A | 0.525 | 0.017 | 0.003 | 322051 | 1.89E-08 | 0.000 | 0.010 | 310537 | 9.02E-01 |
| rs11727676 | T | C | 0.910 | 0.036 | 0.006 | 296401 | 2.55E-08 | -0.011 | 0.017 | 310537 | 8.04E-04 |
| rs11847697[a] | T | C | 0.042 | 0.049 | 0.008 | 306243 | 3.99E-09 | -0.014 | 0.025 | 310537 | 1.68E-03 |
| rs12286929 | G | A | 0.523 | 0.022 | 0.003 | 321903 | 1.31E-12 | 0.016 | 0.010 | 310537 | 1.37E-18 |
| rs12401738 | A | G | 0.352 | 0.021 | 0.003 | 322070 | 1.15E-10 | 0.004 | 0.011 | 310537 | 3.01E-02 |
| rs12429545 | A | G | 0.133 | 0.033 | 0.005 | 312934 | 1.09E-12 | -0.003 | 0.016 | 310537 | 2.15E-01 |
| rs12446632 | G | A | 0.865 | 0.040 | 0.005 | 316758 | 1.48E-18 | -0.005 | 0.015 | 310537 | 3.93E-02 |
| rs12566985[a] | G | A | 0.446 | 0.024 | 0.003 | 319282 | 3.28E-15 | 0.006 | 0.010 | 310537 | 6.04E-04 |
| rs12885454[a] | C | A | 0.642 | 0.021 | 0.003 | 320823 | 1.94E-10 | -0.004 | 0.011 | 310537 | 2.07E-02 |
| rs12940622 | G | A | 0.575 | 0.018 | 0.003 | 322032 | 2.49E-09 | -0.022 | 0.010 | 310537 | 8.00E-33 |
| rs13021737 | G | A | 0.828 | 0.060 | 0.004 | 318287 | 1.11E-50 | 0.005 | 0.014 | 310537 | 4.17E-02 |
| rs13078960 | G | T | 0.196 | 0.030 | 0.004 | 322135 | 1.74E-14 | -0.011 | 0.013 | 310537 | 2.48E-06 |
| rs13107325 | T | C | 0.072 | 0.048 | 0.007 | 321461 | 1.83E-12 | -0.011 | 0.020 | 310537 | 2.57E-03 |
| rs13191362 | A | G | 0.879 | 0.028 | 0.005 | 321902 | 7.34E-09 | -0.002 | 0.016 | 310537 | 4.12E-01 |
| rs1516725 | C | T | 0.872 | 0.045 | 0.005 | 320644 | 1.89E-22 | 0.005 | 0.015 | 310537 | 5.97E-02 |
| rs1528435 | T | C | 0.631 | 0.018 | 0.003 | 321924 | 1.20E-08 | -0.009 | 0.011 | 310537 | 9.40E-07 |
| rs1558902 | A | T | 0.415 | 0.082 | 0.003 | 320073 | 7.51E-153 | 0.008 | 0.011 | 310537 | 2.53E-05 |
| rs16851483 | T | G | 0.066 | 0.048 | 0.008 | 233929 | 3.55E-10 | -0.008 | 0.021 | 310537 | 2.41E-02 |
| rs16951275 | T | C | 0.784 | 0.031 | 0.004 | 322098 | 1.91E-17 | 0.007 | 0.012 | 310537 | 6.99E-04 |
| rs17001654 | G | C | 0.153 | 0.031 | 0.005 | 233722 | 7.76E-09 | 0.015 | 0.015 | 310537 | 1.65E-08 |
| rs17024393 | C | T | 0.040 | 0.066 | 0.009 | 297874 | 7.03E-14 | 0.016 | 0.032 | 310537 | 4.78E-03 |
| rs17094222 | C | T | 0.211 | 0.025 | 0.004 | 321770 | 5.94E-11 | 0.001 | 0.013 | 310537 | 7.17E-01 |
| rs17405819 | T | C | 0.700 | 0.022 | 0.003 | 322085 | 2.07E-11 | 0.002 | 0.011 | 310537 | 4.39E-01 |
| rs17724992 | A | G | 0.746 | 0.019 | 0.004 | 319588 | 3.42E-08 | 0.020 | 0.012 | 310537 | 3.19E-22 |
| rs1808579 | C | T | 0.534 | 0.017 | 0.003 | 322032 | 4.17E-08 | 0.001 | 0.010 | 310537 | 4.37E-01 |
| rs1928295 | T | C | 0.548 | 0.019 | 0.003 | 321979 | 7.91E-10 | 0.001 | 0.010 | 310537 | 5.46E-01 |
| rs2033529 | G | A | 0.293 | 0.019 | 0.003 | 321917 | 1.39E-08 | -0.002 | 0.011 | 310537 | 3.73E-01 |
| rs2033732 | C | T | 0.747 | 0.019 | 0.004 | 321406 | 4.89E-08 | -0.000 | 0.012 | 310537 | 9.30E-01 |
| rs205262 | G | A | 0.273 | 0.022 | 0.004 | 315542 | 1.75E-10 | 0.025 | 0.012 | 310537 | 7.63E-32 |
| rs2075650 | A | G | 0.848 | 0.026 | 0.005 | 308408 | 1.25E-08 | -0.009 | 0.015 | 310537 | 4.83E-04 |
| rs2112347 | T | G | 0.629 | 0.026 | 0.003 | 322019 | 6.19E-17 | 0.013 | 0.011 | 310537 | 2.56E-11 |
| rs2121279 | T | C | 0.152 | 0.025 | 0.004 | 322065 | 2.31E-08 | 0.015 | 0.015 | 310537 | 1.86E-08 |

| | | | | | | | | | | | |
|---|---|---|---|---|---|---|---|---|---|---|---|
| rs2176598 | T | C | 0.251 | 0.020 | 0.004 | 316848 | 2.97E-08 | 0.004 | 0.012 | 310537 | 8.57E-02 |
| rs2207139 | G | A | 0.177 | 0.045 | 0.004 | 322019 | 4.13E-29 | 0.009 | 0.014 | 310537 | 1.68E-04 |
| rs2245368 | C | T | 0.180 | 0.032 | 0.006 | 205675 | 3.19E-08 | 0.008 | 0.014 | 310537 | 1.00E-03 |
| rs2287019[a] | C | T | 0.804 | 0.036 | 0.004 | 300921 | 4.59E-18 | 0.013 | 0.014 | 310537 | 8.26E-08 |
| rs2365389 | C | T | 0.582 | 0.020 | 0.003 | 316768 | 1.63E-10 | -0.007 | 0.011 | 310537 | 3.20E-04 |
| rs2650492 | A | G | 0.303 | 0.021 | 0.004 | 319464 | 1.92E-09 | 0.004 | 0.011 | 310537 | 6.54E-02 |
| rs2820292 | C | A | 0.555 | 0.020 | 0.003 | 321707 | 1.83E-10 | 0.003 | 0.010 | 310537 | 6.24E-02 |
| rs29941 | G | A | 0.669 | 0.018 | 0.003 | 321970 | 2.41E-08 | -0.001 | 0.011 | 310537 | 5.97E-01 |
| rs3101336 | C | T | 0.613 | 0.033 | 0.003 | 316872 | 2.66E-26 | 0.005 | 0.011 | 310537 | 4.49E-03 |
| rs3736485 | A | G | 0.454 | 0.018 | 0.003 | 321398 | 7.41E-09 | 0.017 | 0.010 | 310537 | 1.62E-19 |
| rs3810291[a] | A | G | 0.666 | 0.028 | 0.004 | 296261 | 4.81E-15 | 0.011 | 0.011 | 310537 | 1.46E-08 |
| rs3817334 | T | C | 0.407 | 0.026 | 0.003 | 321959 | 5.15E-17 | -0.004 | 0.010 | 310537 | 5.00E-02 |
| rs3849570 | A | C | 0.359 | 0.019 | 0.003 | 284339 | 2.60E-08 | 0.010 | 0.011 | 310537 | 2.95E-07 |
| rs3888190[a] | A | C | 0.403 | 0.031 | 0.003 | 321930 | 3.14E-23 | 0.013 | 0.011 | 310537 | 1.71E-11 |
| rs4256980 | G | C | 0.646 | 0.021 | 0.003 | 320028 | 2.90E-11 | 0.003 | 0.011 | 310537 | 8.63E-02 |
| rs4740619 | T | C | 0.542 | 0.018 | 0.003 | 321887 | 4.56E-09 | 0.023 | 0.010 | 310537 | 3.67E-36 |
| rs543874 | G | A | 0.193 | 0.048 | 0.004 | 322008 | 2.62E-35 | -0.005 | 0.013 | 310537 | 3.75E-02 |
| rs6477694 | C | T | 0.365 | 0.017 | 0.003 | 322048 | 2.67E-08 | 0.013 | 0.011 | 310537 | 5.67E-12 |
| rs6567160[a] | C | T | 0.236 | 0.056 | 0.004 | 321958 | 3.93E-53 | 0.012 | 0.012 | 310537 | 9.65E-09 |
| rs657452 | A | G | 0.394 | 0.023 | 0.003 | 313651 | 5.48E-13 | 0.014 | 0.011 | 310537 | 9.58E-14 |
| rs6804842 | G | A | 0.575 | 0.019 | 0.003 | 321463 | 2.48E-09 | -0.015 | 0.010 | 310537 | 2.87E-16 |
| rs7138803 | A | G | 0.384 | 0.032 | 0.003 | 322092 | 8.15E-24 | -0.016 | 0.011 | 310537 | 4.60E-16 |
| rs7141420 | T | C | 0.527 | 0.024 | 0.003 | 321970 | 1.23E-14 | 0.004 | 0.010 | 310537 | 2.70E-02 |
| rs7243357 | T | G | 0.812 | 0.022 | 0.004 | 322107 | 3.86E-08 | 0.028 | 0.014 | 310537 | 6.55E-31 |
| rs758747 | T | C | 0.265 | 0.023 | 0.004 | 308688 | 7.47E-10 | -0.009 | 0.012 | 310537 | 5.07E-06 |
| rs7599312 | G | A | 0.724 | 0.022 | 0.003 | 322024 | 1.17E-10 | 0.006 | 0.012 | 310537 | 3.37E-03 |
| rs7899106 | G | A | 0.052 | 0.040 | 0.007 | 321770 | 2.96E-08 | -0.009 | 0.024 | 310537 | 3.19E-02 |
| rs7903146 | C | T | 0.713 | 0.023 | 0.003 | 322130 | 1.11E-11 | -0.000 | 0.011 | 310537 | 9.53E-01 |
| rs9400239 | C | T | 0.688 | 0.019 | 0.003 | 321988 | 1.61E-08 | 0.017 | 0.011 | 310537 | 7.57E-18 |
| rs9925964[a] | A | G | 0.620 | 0.019 | 0.003 | 318385 | 8.11E-10 | 0.011 | 0.011 | 310537 | 9.47E-09 |

BMI = body mass index, EA = effect allele, EAF = effect allele frequency, LD = linkage disequilibrium, MR = Mendelian randomization, OA = other allele, SE = standard error

a) These 12 SNPs exceeded the LD threshold ($R^2$<0.001) and were excluded from the two-sample summary MR analysis

b) The effect allele (EA) is the BMI increasing allele

c) Coefficients are given per BMI SD (4.6 kg/$m^2$)

d) Coefficients are given on the logarithmic scale, obtained from a quasi-Poisson regression of hospital admission count on the relevant SNP, using person-years on study as an offset and adjusting for age, sex and the first 40 genetic principle components

**Table S4.** WHR-SNP and hospital admission count-SNP associations as used for the univariate two-sample MR analysis of the effect of WHR on yearly hospital admission rate. The WHR-SNP association coefficients were used as external weights for the genetic risk score in the one-sample MR analysis.

| | | | | **WHR-SNP associations (European ancestry GIANT consortium meta-analysis, Shungin *et al.* (2015)** | | | | **Hospital admission count – SNP associations (UK Biobank)** | | | |
|---|---|---|---|---|---|---|---|---|---|---|---|
| **SNP** | **EA[b]** | **OA** | **EAF** | **β[c]** | **SE** | ***N*** | ***P*** | **β[d]** | **SE** | ***N*** | ***P*** |
| rs1011731[a] | G | A | 0.427 | 0.019 | 0.003 | 212094 | 1.07E-08 | -0.010 | 0.010 | 310537 | 7.08E-08 |
| rs10195252 | T | C | 0.590 | 0.020 | 0.003 | 211907 | 2.57E-09 | 0.005 | 0.011 | 310537 | 9.27E-03 |
| rs10245353 | A | C | 0.200 | 0.027 | 0.004 | 212151 | 1.57E-10 | 0.006 | 0.013 | 310537 | 9.64E-03 |
| rs1045241 | C | T | 0.714 | 0.015 | 0.004 | 212012 | 5.76E-05 | -0.008 | 0.012 | 310537 | 8.74E-05 |
| rs10804591 | A | C | 0.793 | 0.021 | 0.004 | 212108 | 2.09E-07 | -0.005 | 0.013 | 310537 | 1.63E-02 |
| rs11048470 | T | G | 0.277 | 0.025 | 0.004 | 212159 | 6.33E-12 | -0.001 | 0.012 | 310537 | 6.45E-01 |
| rs1121980 | A | G | 0.433 | 0.043 | 0.003 | 211970 | 1.33E-38 | 0.011 | 0.010 | 310537 | 5.44E-09 |
| rs11663816 | C | T | 0.265 | 0.025 | 0.004 | 212104 | 2.65E-11 | 0.006 | 0.012 | 310537 | 4.99E-03 |
| rs11989744 | C | T | 0.760 | 0.021 | 0.005 | 144546 | 1.30E-05 | 0.007 | 0.012 | 310537 | 5.43E-04 |
| rs12549058 | G | T | 0.084 | 0.040 | 0.006 | 212028 | 3.17E-10 | -0.004 | 0.022 | 310537 | 3.19E-01 |
| rs1294421 | G | T | 0.620 | 0.026 | 0.003 | 212054 | 6.93E-14 | -0.007 | 0.011 | 310537 | 4.15E-04 |
| rs1358980 | T | C | 0.470 | 0.027 | 0.004 | 211037 | 1.98E-14 | 0.014 | 0.010 | 310537 | 1.48E-13 |
| rs1394461 | C | G | 0.251 | 0.017 | 0.005 | 144349 | 4.68E-04 | 0.005 | 0.013 | 310537 | 1.33E-02 |
| rs1440372 | C | T | 0.710 | 0.021 | 0.004 | 210387 | 7.59E-09 | 0.014 | 0.012 | 310537 | 4.20E-11 |
| rs1443512 | A | C | 0.235 | 0.026 | 0.004 | 212153 | 2.76E-11 | 0.001 | 0.013 | 310537 | 7.11E-01 |
| rs1515108 | C | T | 0.377 | 0.007 | 0.003 | 212094 | 3.13E-02 | -0.020 | 0.011 | 310537 | 8.41E-26 |
| rs1569135 | A | G | 0.529 | 0.024 | 0.003 | 212086 | 1.00E-12 | 0.012 | 0.010 | 310537 | 2.61E-10 |
| rs16996700 | T | C | 0.730 | 0.021 | 0.004 | 212159 | 1.60E-08 | 0.014 | 0.011 | 310537 | 8.92E-12 |
| rs17109256 | A | G | 0.219 | 0.023 | 0.004 | 208422 | 3.02E-08 | 0.005 | 0.013 | 310537 | 2.67E-02 |
| rs17451107 | T | C | 0.613 | 0.023 | 0.004 | 211586 | 3.50E-11 | -0.005 | 0.011 | 310537 | 1.26E-02 |
| rs17819328 | G | T | 0.432 | 0.016 | 0.004 | 211496 | 2.29E-06 | 0.029 | 0.010 | 310537 | 2.35E-55 |
| rs2075650 | A | G | 0.848 | 0.029 | 0.005 | 206613 | 6.43E-09 | -0.009 | 0.015 | 310537 | 4.83E-04 |
| rs2179129 | A | G | 0.590 | 0.021 | 0.003 | 212181 | 1.24E-09 | 0.016 | 0.011 | 310537 | 1.54E-18 |
| rs2287019[a] | C | T | 0.802 | 0.026 | 0.005 | 199713 | 4.34E-09 | 0.013 | 0.014 | 310537 | 8.26E-08 |
| rs2765539 | T | C | 0.735 | 0.027 | 0.004 | 212176 | 1.08E-12 | 0.001 | 0.012 | 310537 | 6.31E-01 |
| rs319564 | C | T | 0.450 | 0.014 | 0.003 | 212137 | 3.42E-05 | 0.012 | 0.010 | 310537 | 4.00E-10 |
| rs3786897 | G | A | 0.417 | 0.022 | 0.003 | 212009 | 3.95E-11 | -0.002 | 0.010 | 310537 | 2.46E-01 |
| rs4471313 | T | G | 0.720 | 0.020 | 0.004 | 180767 | 6.92E-07 | 0.025 | 0.012 | 310537 | 6.19E-33 |
| rs459193 | A | G | 0.264 | 0.026 | 0.004 | 212101 | 6.02E-12 | -0.008 | 0.013 | 310537 | 3.67E-04 |
| rs4640244[a] | G | A | 0.396 | 0.021 | 0.004 | 198799 | 3.11E-08 | 0.013 | 0.011 | 310537 | 1.06E-11 |
| rs4646404 | G | A | 0.661 | 0.020 | 0.004 | 201330 | 3.81E-07 | -0.002 | 0.011 | 310537 | 3.19E-01 |
| rs4715208[a] | G | A | 0.740 | 0.019 | 0.004 | 212191 | 7.91E-07 | -0.004 | 0.012 | 310537 | 7.70E-02 |
| rs4846565 | G | A | 0.672 | 0.023 | 0.004 | 212157 | 4.75E-11 | 0.009 | 0.011 | 310537 | 5.30E-06 |
| rs4929927 | G | A | 0.646 | 0.020 | 0.003 | 212152 | 7.60E-09 | 0.004 | 0.011 | 310537 | 4.70E-02 |
| rs7801581[a] | T | C | 0.244 | 0.023 | 0.004 | 198342 | 4.95E-08 | -0.022 | 0.012 | 310537 | 7.73E-24 |
| rs863750 | T | C | 0.593 | 0.016 | 0.003 | 212134 | 1.56E-06 | 0.003 | 0.011 | 310537 | 1.33E-01 |
| rs929641 | A | G | 0.587 | 0.020 | 0.003 | 212102 | 4.25E-09 | -0.015 | 0.011 | 310537 | 3.58E-16 |
| rs9491696 | G | C | 0.482 | 0.038 | 0.003 | 211988 | 4.88E-30 | 0.005 | 0.010 | 310537 | 1.37E-02 |
| rs9860730 | A | G | 0.703 | 0.023 | 0.004 | 212062 | 2.84E-10 | 0.005 | 0.011 | 310537 | 1.14E-02 |

EA = effect allele, EAF = effect allele frequency, LD = linkage disequilibrium, MR = Mendelian randomization, OA = other allele, SE = standard error, WHR = waist-hip-ratio

a) These 5 SNPs exceeded the LD threshold ($R^2$<0.001) and were excluded from the two-sample summary MR analysis

b) The effect allele (EA) is the WHR increasing allele

c) Coefficients are given per WHR SD (0.07)

d) Coefficients are given on the logarithmic scale, obtained from a quasi-Poisson regression of hospital admission count on the relevant SNP, using person-years on study as an offset and adjusting for age, sex and the first 40 genetic principle components

**Table S5.** WHRadjBMI-SNP and hospital admission count-SNP associations as used for the univariate two-sample MR analysis of the effect of WHRadjBMI on yearly hospital admission rate. The WHRadjBMI-SNP association coefficients were used as external weights for the genetic risk score in the one-sample MR analysis

| | | | | **WHRadjBMI-SNP associations (European ancestry GIANT consortium meta-analysis, Shungin *et al.* (2015)** | | | | **Hospital admission count – SNP associations (UK Biobank)** | | | |
|---|---|---|---|---|---|---|---|---|---|---|---|
| **SNP** | **EA**[b] | **OA** | **EAF** | **β**[c] | **SE** | ***N*** | ***P*** | **β**[d] | **SE** | ***N*** | ***P*** |
| rs10195252 | T | C | 0.587 | 0.031 | 0.004 | 142102 | 1.03E-15 | 0.005 | 0.011 | 310537 | 9.27E-03 |
| rs10245353 | A | C | 0.196 | 0.037 | 0.005 | 142708 | 6.88E-14 | 0.006 | 0.013 | 310537 | 9.64E-03 |
| rs1045241 | C | T | 0.714 | 0.022 | 0.004 | 142400 | 3.18E-07 | -0.008 | 0.012 | 310537 | 8.74E-05 |
| rs10804591 | A | C | 0.794 | 0.025 | 0.005 | 142653 | 1.93E-07 | -0.005 | 0.013 | 310537 | 1.63E-02 |
| rs10842707 | T | C | 0.227 | 0.036 | 0.005 | 142708 | 4.39E-15 | -0.003 | 0.013 | 310537 | 1.53E-01 |
| rs10919388 | C | A | 0.721 | 0.025 | 0.004 | 142721 | 5.33E-09 | 0.025 | 0.012 | 310537 | 8.67E-33 |
| rs10991437 | A | C | 0.117 | 0.033 | 0.006 | 142661 | 9.74E-08 | -0.003 | 0.016 | 310537 | 2.41E-01 |
| rs11231693 | A | G | 0.061 | 0.048 | 0.009 | 130856 | 8.20E-08 | 0.008 | 0.023 | 310537 | 3.54E-02 |
| rs12454712 | T | C | 0.615 | 0.017 | 0.006 | 102489 | 3.26E-03 | 0.004 | 0.011 | 310537 | 1.84E-02 |
| rs12608504 | A | G | 0.354 | 0.020 | 0.004 | 142678 | 1.20E-06 | 0.010 | 0.011 | 310537 | 6.65E-07 |
| rs12679556 | G | T | 0.245 | 0.024 | 0.005 | 142669 | 1.12E-07 | 0.006 | 0.012 | 310537 | 3.20E-03 |
| rs1294410 | C | T | 0.633 | 0.034 | 0.004 | 142548 | 1.42E-17 | -0.003 | 0.011 | 310537 | 6.55E-02 |
| rs1358980 | T | C | 0.461 | 0.039 | 0.004 | 139579 | 3.01E-20 | 0.014 | 0.010 | 310537 | 1.48E-13 |
| rs1385167 | G | A | 0.142 | 0.032 | 0.006 | 139368 | 9.40E-09 | -0.028 | 0.015 | 310537 | 7.87E-25 |
| rs1440372 | C | T | 0.705 | 0.023 | 0.004 | 141188 | 7.84E-08 | 0.014 | 0.012 | 310537 | 4.20E-11 |
| rs1443512 | A | C | 0.233 | 0.031 | 0.005 | 142694 | 5.38E-12 | 0.001 | 0.013 | 310537 | 7.11E-01 |
| rs1569135 | A | G | 0.528 | 0.020 | 0.004 | 142674 | 3.58E-07 | 0.012 | 0.010 | 310537 | 2.61E-10 |
| rs17451107 | T | C | 0.614 | 0.027 | 0.004 | 140959 | 1.48E-10 | -0.005 | 0.011 | 310537 | 1.26E-02 |
| rs1776897 | G | T | 0.082 | 0.041 | 0.008 | 110603 | 5.50E-07 | 0.011 | 0.018 | 310537 | 1.24E-03 |
| rs17819328 | G | T | 0.431 | 0.022 | 0.004 | 141503 | 5.82E-08 | 0.029 | 0.010 | 310537 | 2.35E-55 |
| rs1936805 | T | C | 0.509 | 0.042 | 0.004 | 142540 | 6.13E-28 | 0.006 | 0.010 | 310537 | 1.69E-03 |
| rs224333 | G | A | 0.634 | 0.022 | 0.004 | 142206 | 2.76E-07 | 0.002 | 0.011 | 310537 | 2.90E-01 |
| rs2276824 | C | G | 0.433 | 0.022 | 0.004 | 141626 | 7.12E-08 | -0.007 | 0.010 | 310537 | 6.07E-05 |
| rs2294239 | A | G | 0.586 | 0.028 | 0.004 | 142140 | 1.86E-12 | 0.016 | 0.010 | 310537 | 1.26E-17 |
| rs2371767 | G | C | 0.723 | 0.034 | 0.005 | 130378 | 9.49E-14 | 0.000 | 0.012 | 310537 | 8.52E-01 |
| rs2645294 | T | C | 0.575 | 0.034 | 0.004 | 142526 | 4.42E-18 | 0.002 | 0.010 | 310537 | 3.07E-01 |
| rs2820443 | T | C | 0.714 | 0.041 | 0.004 | 142663 | 1.50E-21 | 0.002 | 0.011 | 310537 | 2.86E-01 |
| rs2925979 | T | C | 0.305 | 0.018 | 0.004 | 140533 | 1.44E-05 | 0.010 | 0.011 | 310537 | 2.29E-07 |
| rs303084 | A | G | 0.798 | 0.024 | 0.005 | 142629 | 5.30E-07 | -0.006 | 0.013 | 310537 | 5.39E-03 |
| rs3805389 | A | G | 0.279 | 0.018 | 0.004 | 141909 | 4.16E-05 | 0.011 | 0.012 | 310537 | 3.00E-08 |
| rs4081724 | G | A | 0.856 | 0.031 | 0.006 | 140159 | 1.25E-07 | 0.018 | 0.015 | 310537 | 1.40E-11 |
| rs4646404 | G | A | 0.665 | 0.026 | 0.005 | 131007 | 3.46E-08 | -0.002 | 0.011 | 310537 | 3.19E-01 |
| rs4765219 | C | A | 0.667 | 0.030 | 0.004 | 142549 | 5.99E-14 | 0.005 | 0.011 | 310537 | 1.02E-02 |
| rs6090583 | A | G | 0.474 | 0.020 | 0.004 | 142152 | 2.47E-07 | 0.028 | 0.010 | 310537 | 1.56E-50 |
| rs6556301[a] | T | G | 0.356 | 0.022 | 0.004 | 140524 | 2.60E-07 | -0.006 | 0.011 | 310537 | 3.89E-03 |
| rs714515[a] | G | A | 0.428 | 0.031 | 0.004 | 142435 | 6.72E-16 | -0.010 | 0.010 | 310537 | 2.89E-08 |
| rs7705502 | A | G | 0.320 | 0.023 | 0.004 | 142668 | 2.38E-08 | -0.021 | 0.011 | 310537 | 9.98E-26 |
| rs7759742 | A | T | 0.505 | 0.024 | 0.004 | 140972 | 1.50E-09 | 0.013 | 0.010 | 310537 | 6.67E-13 |
| rs7801581[a] | T | C | 0.242 | 0.027 | 0.005 | 129075 | 5.02E-08 | -0.022 | 0.012 | 310537 | 7.73E-24 |
| rs7830933 | A | G | 0.766 | 0.021 | 0.005 | 142458 | 4.73E-06 | 0.013 | 0.012 | 310537 | 7.00E-10 |
| rs7917772 | A | G | 0.620 | 0.017 | 0.004 | 142345 | 3.20E-05 | 0.003 | 0.011 | 310537 | 9.90E-02 |
| rs8030605 | A | G | 0.150 | 0.029 | 0.006 | 141175 | 1.66E-06 | 0.003 | 0.016 | 310537 | 2.47E-01 |
| rs8042543 | C | T | 0.795 | 0.024 | 0.005 | 141009 | 3.45E-06 | -0.003 | 0.013 | 310537 | 1.36E-01 |
| rs8066985 | A | G | 0.507 | 0.019 | 0.004 | 142684 | 1.28E-06 | 0.001 | 0.010 | 310537 | 8.06E-01 |
| rs905938 | T | C | 0.738 | 0.030 | 0.005 | 140583 | 1.73E-10 | -0.013 | 0.012 | 310537 | 8.45E-10 |
| rs9687846 | A | G | 0.186 | 0.027 | 0.005 | 142400 | 8.28E-08 | -0.002 | 0.013 | 310537 | 4.01E-01 |

| | | | | | | | | | | | |
|---|---|---|---|---|---|---|---|---|---|---|---|
| rs979012 | T | C | 0.347 | 0.026 | 0.004 | 142646 | 4.22E-10 | 0.013 | 0.011 | 310537 | 5.13E-12 |
| rs9991328 | T | C | 0.480 | 0.019 | 0.004 | 142624 | 5.87E-07 | -0.014 | 0.010 | 310537 | 4.62E-14 |

BMI = body mass index, EA = effect allele, EAF = effect allele frequency, LD = linkage disequilibrium, MR = Mendelian randomization, OA = other allele, SE = standard error, WHRadjBMI = waist-hip-ratio adjusted for BMI

a) These 3 SNPs exceeded the LD threshold ($R^2$<0.001) and were excluded from the two-sample summary MR analysis

b) The effect allele (EA) is the WHR increasing allele

c) Coefficients are given per WHR SD (0.07)

d) Coefficients are given on the logarithmic scale, obtained from a quasi-Poisson regression of hospital admission count on the relevant SNP, using person-years on study as an offset and adjusting for age, sex and the first 40 PCAs

**Table S6.** BMI-SNP, WHR-SNP and hospital admission count-SNP associations as used for the multivariable two-sample MR analysis of the effect of BMI and WHR on yearly hospital admission rate. Given are the 70 SNPs remaining after LD correction ($R^2$<0.001) to the joint set of BMI and WHR SNPs.

| | | | | **BMI-SNP associations (European ancestry GIANT consortium meta-analysis, Locke *et al.* (2016)** | | | | **WHR-SNP associations (European ancestry GIANT consortium meta-analysis, Shungin *et al.* (2015)** | | | | **Hospital admission count – SNP associations (UK Biobank, *N*=310537)** | | |
|---|---|---|---|---|---|---|---|---|---|---|---|---|---|---|
| **SNP** | **EA** | **OA** | **EAF** | **β[a]** | **SE** | ***P*** | ***N*** | **β[a]** | **SE** | ***P*** | ***N*** | **β[b]** | **SE** | ***P*** |
| rs1000940 | G | A | 0.225 | 0.019 | 0.003 | 1.28E-08 | 321836 | 0.008 | 0.004 | 2.30E-02 | 211915 | -0.012 | 0.011 | 9.25E-09 |
| rs1011731 | G | A | 0.458 | -0.006 | 0.003 | 3.88E-02 | 321942 | 0.019 | 0.003 | 1.10E-08 | 212094 | -0.010 | 0.010 | 7.08E-08 |
| rs10132280 | A | C | 0.333 | -0.023 | 0.003 | 1.14E-11 | 321797 | -0.012 | 0.004 | 8.40E-04 | 212105 | 0.003 | 0.011 | 1.76E-01 |
| rs1016287 | T | C | 0.325 | 0.023 | 0.003 | 2.25E-11 | 321969 | 0.013 | 0.004 | 6.20E-04 | 212119 | 0.015 | 0.011 | 9.98E-14 |
| rs10182181 | A | G | 0.500 | -0.031 | 0.003 | 8.78E-24 | 321759 | -0.005 | 0.003 | 1.60E-01 | 211882 | -0.009 | 0.010 | 1.11E-06 |
| rs10245353 | A | C | 0.183 | -0.002 | 0.004 | 5.88E-01 | 322088 | 0.027 | 0.004 | 1.60E-10 | 212151 | 0.006 | 0.013 | 9.64E-03 |
| rs10733682 | A | G | 0.425 | 0.017 | 0.003 | 1.83E-08 | 320727 | 0.010 | 0.003 | 2.90E-03 | 211864 | -0.003 | 0.010 | 1.24E-01 |
| rs10938397 | A | G | 0.567 | -0.040 | 0.003 | 3.20E-38 | 320955 | -0.018 | 0.003 | 2.40E-07 | 211843 | -0.004 | 0.010 | 5.44E-02 |
| rs10968576 | G | A | 0.292 | 0.025 | 0.003 | 6.61E-14 | 322061 | 0.017 | 0.004 | 2.40E-06 | 212162 | -0.001 | 0.011 | 6.46E-01 |
| rs11030104 | A | G | 0.800 | 0.041 | 0.004 | 5.56E-28 | 322103 | 0.020 | 0.004 | 8.50E-07 | 212160 | 0.014 | 0.013 | 1.43E-09 |
| rs11048470 | T | G | 0.233 | -0.009 | 0.003 | 6.35E-03 | 322043 | 0.025 | 0.004 | 6.30E-12 | 212159 | -0.001 | 0.012 | 6.45E-01 |
| rs11165643 | C | T | 0.425 | -0.022 | 0.003 | 2.07E-12 | 320730 | -0.009 | 0.003 | 6.60E-03 | 212148 | 0.012 | 0.011 | 5.15E-10 |
| rs1167827 | A | G | 0.458 | -0.020 | 0.003 | 6.33E-10 | 306238 | -0.006 | 0.004 | 8.00E-02 | 204123 | -0.004 | 0.010 | 4.97E-02 |
| rs11727676 | C | T | 0.075 | 0.036 | 0.006 | 2.55E-08 | 296401 | 0.005 | 0.007 | 4.30E-01 | 191737 | -0.011 | 0.017 | 8.04E-04 |
| rs12286929 | G | A | 0.433 | 0.022 | 0.003 | 1.31E-12 | 321903 | 0.010 | 0.003 | 4.10E-03 | 212086 | 0.016 | 0.010 | 1.37E-18 |
| rs12429545 | G | A | 0.900 | 0.033 | 0.005 | 1.09E-12 | 312934 | -0.015 | 0.005 | 2.90E-03 | 203113 | -0.003 | 0.016 | 2.15E-01 |
| rs12940622 | A | G | 0.458 | -0.018 | 0.003 | 2.49E-09 | 322032 | -0.007 | 0.003 | 2.60E-02 | 212119 | -0.007 | 0.011 | 8.00E-33 |
| rs1294421 | G | T | 0.600 | -0.005 | 0.003 | 1.42E-01 | 321751 | 0.025 | 0.003 | 6.90E-14 | 212054 | 0.005 | 0.014 | 4.15E-04 |
| rs13021737 | A | G | 0.125 | -0.060 | 0.004 | 1.11E-50 | 318287 | -0.023 | 0.004 | 1.90E-07 | 209902 | -0.011 | 0.013 | 4.17E-02 |
| rs13078960 | T | G | 0.817 | -0.030 | 0.004 | 1.74E-14 | 322135 | -0.010 | 0.004 | 2.10E-02 | 212190 | -0.011 | 0.020 | 2.48E-06 |
| rs13107325 | C | T | 0.883 | -0.048 | 0.007 | 1.82E-12 | 321461 | 0.000 | 0.007 | 1.00E+00 | 211675 | -0.002 | 0.016 | 2.57E-03 |
| rs13191362 | A | G | 0.800 | 0.028 | 0.005 | 7.34E-09 | 321902 | 0.018 | 0.005 | 6.00E-04 | 212035 | 0.005 | 0.015 | 4.12E-01 |
| rs1516725 | T | C | 0.092 | -0.045 | 0.005 | 1.89E-22 | 320644 | -0.014 | 0.005 | 4.40E-03 | 210725 | 0.012 | 0.010 | 5.97E-02 |
| rs1569135 | A | G | 0.533 | 0.011 | 0.003 | 5.24E-04 | 322000 | 0.024 | 0.003 | 1.00E-12 | 212086 | -0.008 | 0.021 | 2.61E-10 |
| rs16851483 | G | T | 0.908 | -0.048 | 0.008 | 3.55E-10 | 233929 | -0.019 | 0.008 | 2.20E-02 | 144591 | 0.007 | 0.012 | 2.41E-02 |
| rs16951275 | C | T | 0.225 | -0.031 | 0.004 | 1.91E-17 | 322098 | -0.016 | 0.004 | 5.00E-05 | 212172 | 0.014 | 0.011 | 6.99E-04 |
| rs16996700 | T | C | 0.700 | 0.018 | 0.003 | 1.74E-07 | 322063 | 0.021 | 0.004 | 1.60E-08 | 212159 | 0.015 | 0.015 | 8.92E-12 |
| rs17001654 | C | G | 0.842 | -0.031 | 0.005 | 7.76E-09 | 233722 | -0.010 | 0.006 | 9.30E-02 | 144378 | 0.016 | 0.032 | 1.65E-08 |
| rs17024393 | C | T | 0.042 | 0.066 | 0.009 | 7.03E-14 | 297874 | 0.029 | 0.009 | 1.80E-03 | 189151 | 0.001 | 0.013 | 4.78E-03 |
| rs17094222 | C | T | 0.208 | 0.025 | 0.004 | 5.94E-11 | 321770 | 0.013 | 0.004 | 2.00E-03 | 212078 | 0.002 | 0.011 | 7.17E-01 |
| rs17405819 | C | T | 0.367 | -0.022 | 0.003 | 2.07E-11 | 322085 | -0.002 | 0.004 | 6.80E-01 | 212166 | -0.005 | 0.011 | 4.39E-01 |
| rs17451107 | T | C | 0.625 | 0.005 | 0.003 | 1.58E-01 | 320725 | 0.023 | 0.004 | 3.50E-11 | 211586 | 0.020 | 0.012 | 1.26E-02 |
| rs17724992 | A | G | 0.692 | 0.019 | 0.004 | 3.42E-08 | 319588 | 0.019 | 0.004 | 9.30E-07 | 210660 | 0.001 | 0.010 | 3.19E-22 |
| rs1808579 | T | C | 0.475 | -0.017 | 0.003 | 4.17E-08 | 322032 | -0.004 | 0.003 | 1.80E-01 | 212105 | 0.001 | 0.010 | 4.37E-01 |
| rs1928295 | C | T | 0.425 | -0.019 | 0.003 | 7.91E-10 | 321979 | -0.012 | 0.003 | 3.90E-04 | 212089 | -0.002 | 0.011 | 5.46E-01 |
| rs2033529 | G | A | 0.258 | 0.019 | 0.003 | 1.39E-08 | 321917 | 0.016 | 0.004 | 6.00E-06 | 212146 | -0.000 | 0.012 | 3.73E-01 |
| rs2033732 | C | T | 0.758 | 0.019 | 0.004 | 4.89E-08 | 321406 | 0.009 | 0.004 | 1.80E-02 | 211985 | 0.025 | 0.012 | 9.30E-01 |
| rs205262 | A | G | 0.733 | -0.022 | 0.004 | 1.75E-10 | 315542 | 0.000 | 0.004 | 9.80E-01 | 205608 | 0.013 | 0.011 | 7.63E-32 |
| rs2112347 | G | T | 0.375 | -0.026 | 0.003 | 6.19E-17 | 322019 | -0.013 | 0.003 | 2.50E-04 | 212132 | 0.015 | 0.015 | 2.56E-11 |
| rs2121279 | T | C | 0.117 | 0.024 | 0.004 | 2.31E-08 | 322065 | 0.010 | 0.005 | 4.20E-02 | 212154 | 0.004 | 0.012 | 1.86E-08 |
| rs2176598 | T | C | 0.200 | 0.020 | 0.004 | 2.97E-08 | 316848 | 0.016 | 0.004 | 3.50E-05 | 206987 | 0.016 | 0.011 | 8.57E-02 |
| rs2179129 | A | G | 0.550 | -0.002 | 0.003 | 5.87E-01 | 321616 | 0.021 | 0.003 | 1.20E-09 | 212181 | 0.009 | 0.014 | 1.54E-18 |
| rs2207139 | G | A | 0.100 | 0.045 | 0.004 | 4.13E-29 | 322019 | 0.025 | 0.004 | 1.40E-08 | 212163 | 0.008 | 0.014 | 1.68E-04 |
| rs2245368 | T | C | 0.758 | -0.032 | 0.006 | 3.19E-08 | 205675 | -0.009 | 0.006 | 1.30E-01 | 133817 | 0.013 | 0.014 | 1.00E-03 |
| rs2287019 | C | T | 0.850 | 0.036 | 0.004 | 4.58E-18 | 300921 | 0.026 | 0.004 | 4.30E-09 | 199713 | 0.013 | 0.014 | 8.26E-08 |

| | | | | | | | | | | | | | |
|---|---|---|---|---|---|---|---|---|---|---|---|---|---|
| rs2365389 | C | T | 0.658 | 0.020 | 0.003 | 1.63E-10 | 316768 | 0.008 | 0.003 | 2.20E-02 | 208487 | -0.007 | 0.011 | 3.20E-04 |
| rs2820292 | A | C | 0.492 | -0.020 | 0.003 | 1.83E-10 | 321707 | -0.011 | 0.003 | 7.80E-04 | 212043 | 0.003 | 0.010 | 6.24E-02 |
| rs3101336 | T | C | 0.351 | -0.033 | 0.003 | 2.66E-26 | 316872 | -0.016 | 0.003 | 3.60E-06 | 206980 | 0.005 | 0.011 | 4.49E-03 |
| rs3736485 | A | G | 0.425 | 0.018 | 0.003 | 7.41E-09 | 321398 | 0.015 | 0.003 | 6.60E-06 | 212010 | 0.017 | 0.010 | 1.62E-19 |
| rs3786897 | G | A | 0.408 | 0.007 | 0.003 | 3.43E-02 | 321528 | 0.022 | 0.003 | 4.00E-11 | 212009 | -0.002 | 0.010 | 2.46E-01 |
| rs3817334 | C | T | 0.550 | -0.026 | 0.003 | 5.14E-17 | 321959 | -0.009 | 0.003 | 6.10E-03 | 212046 | -0.004 | 0.010 | 5.00E-02 |
| rs3849570 | A | C | 0.367 | 0.019 | 0.003 | 2.60E-08 | 284339 | 0.011 | 0.004 | 2.20E-03 | 182853 | 0.010 | 0.011 | 2.95E-07 |
| rs3888190 | A | C | 0.358 | 0.031 | 0.003 | 3.14E-23 | 321930 | 0.016 | 0.003 | 1.40E-06 | 212068 | 0.013 | 0.011 | 1.71E-11 |
| rs4256980 | G | C | 0.725 | 0.021 | 0.003 | 2.90E-11 | 320028 | 0.019 | 0.003 | 5.30E-08 | 210672 | 0.003 | 0.011 | 8.63E-02 |
| rs459193 | A | G | 0.217 | 0.007 | 0.004 | 4.29E-02 | 321858 | 0.026 | 0.004 | 6.00E-12 | 212101 | -0.008 | 0.013 | 3.67E-04 |
| rs4640244 | G | A | 0.375 | 0.016 | 0.003 | 4.57E-06 | 305292 | 0.021 | 0.004 | 3.10E-08 | 198799 | 0.013 | 0.011 | 1.06E-11 |
| rs4740619 | T | C | 0.533 | 0.018 | 0.003 | 4.56E-09 | 321887 | 0.011 | 0.003 | 1.10E-03 | 212076 | 0.023 | 0.010 | 3.67E-36 |
| rs543874 | G | A | 0.267 | 0.048 | 0.004 | 2.62E-35 | 322008 | 0.020 | 0.004 | 2.00E-06 | 212160 | -0.005 | 0.013 | 3.75E-02 |
| rs6477694 | C | T | 0.358 | 0.017 | 0.003 | 2.67E-08 | 322048 | 0.015 | 0.003 | 1.50E-05 | 212114 | 0.013 | 0.011 | 5.67E-12 |
| rs6567160 | C | T | 0.283 | 0.056 | 0.004 | 3.93E-53 | 321958 | 0.025 | 0.004 | 5.90E-10 | 212147 | 0.012 | 0.012 | 9.65E-09 |
| rs657452 | A | G | 0.417 | 0.023 | 0.003 | 5.48E-13 | 313651 | 0.014 | 0.004 | 4.20E-05 | 204624 | 0.014 | 0.011 | 9.58E-14 |
| rs6804842 | A | G | 0.425 | -0.018 | 0.003 | 2.48E-09 | 321463 | -0.006 | 0.003 | 5.80E-02 | 212015 | -0.015 | 0.010 | 2.87E-16 |
| rs7138803 | G | A | 0.558 | -0.032 | 0.003 | 8.15E-24 | 322092 | -0.014 | 0.004 | 3.50E-05 | 212167 | -0.016 | 0.011 | 4.60E-16 |
| rs7141420 | T | C | 0.617 | 0.024 | 0.003 | 1.23E-14 | 321970 | 0.017 | 0.003 | 5.10E-07 | 212098 | 0.004 | 0.010 | 2.70E-02 |
| rs758747 | C | T | 0.733 | -0.022 | 0.004 | 7.47E-10 | 308688 | -0.007 | 0.004 | 6.30E-02 | 200285 | -0.009 | 0.012 | 5.07E-06 |
| rs7599312 | G | A | 0.708 | 0.022 | 0.003 | 1.17E-10 | 322024 | 0.009 | 0.004 | 1.70E-02 | 212134 | 0.006 | 0.012 | 3.37E-03 |
| rs7899106 | A | G | 0.950 | -0.040 | 0.007 | 2.96E-08 | 321770 | -0.011 | 0.008 | 1.40E-01 | 212004 | -0.009 | 0.024 | 3.19E-02 |
| rs7903146 | T | C | 0.250 | -0.023 | 0.003 | 1.11E-11 | 322130 | -0.003 | 0.004 | 4.30E-01 | 212185 | -0.000 | 0.011 | 9.53E-01 |
| rs929641 | A | G | 0.617 | 0.017 | 0.003 | 1.11E-07 | 322004 | 0.020 | 0.003 | 4.20E-09 | 212102 | -0.015 | 0.011 | 3.58E-16 |
| rs9400239 | C | T | 0.700 | 0.019 | 0.003 | 1.61E-08 | 321988 | 0.015 | 0.004 | 2.10E-05 | 212123 | 0.017 | 0.011 | 7.57E-18 |

BMI = body mass index, EA = effect allele, EAF = effect allele frequency, LD = linkage disequilibrium, MR = Mendelian randomization, OA = other allele, SE = standard error, WHR = waist-hip-ratio

a) Coefficients are given per SD unit, 4.6 kg/$m^2$ and 0.07, for BMI and WHR, respectively, consistent with the Locke (2016) and Shungin (2015) GWAS meta-analyses. For the analyses the coefficients were used on the SD scale for both exposures and rescaled to whole units for BMI and 0.1 units for WHR.

b) Coefficients are given on the logarithmic scale, obtained from a quasi-Poisson regression of hospital admission count on the relevant SNP, using person-years on study as an offset and adjusting for age, sex and the first 40 PCAs

**Figure S1**. Plots for two-sample MR analysis of BMI effect (per SD, with $SD_{BMI}$=4.6) on yearly hospital admission rate in UK Biobank participants of White British ancestry. Effects are shown on the log(rate) scale. A) Cochran's Q is plotted against Rucker's Q for each SNP, as calculated in a leave-one-out analysis. Outliers have been visually identified and are labeled. Q-statistics for the model with the full set of 64 SNPs (red) and with outliers removed (61 SNPs, blue) are shown. B) SNP effect on yearly hospital admission count is plotted against the SNP effect on BMI. Fitted lines are displayed for the random effects MR-Egger, penalized weighted median, weighted mode and random effects exact weight IVW models. The outliers identified in plot *A* are shown in blue. * denotes a significant slope.

**BMI**

**A**

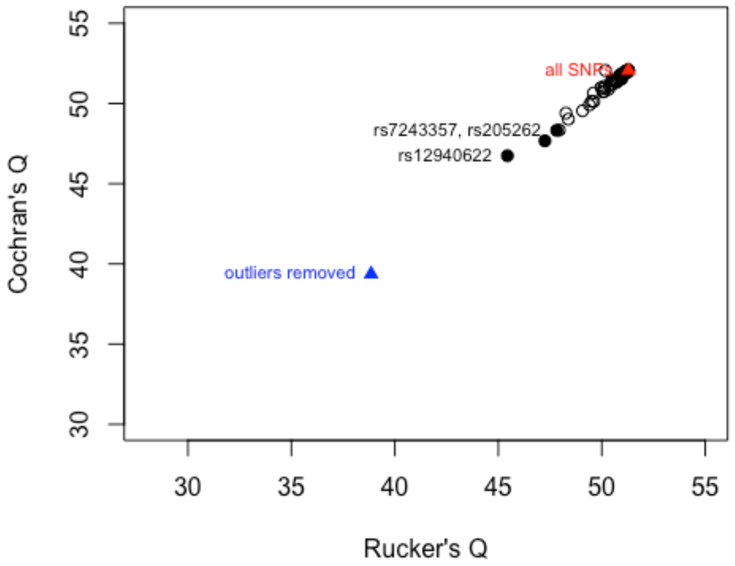


**B**

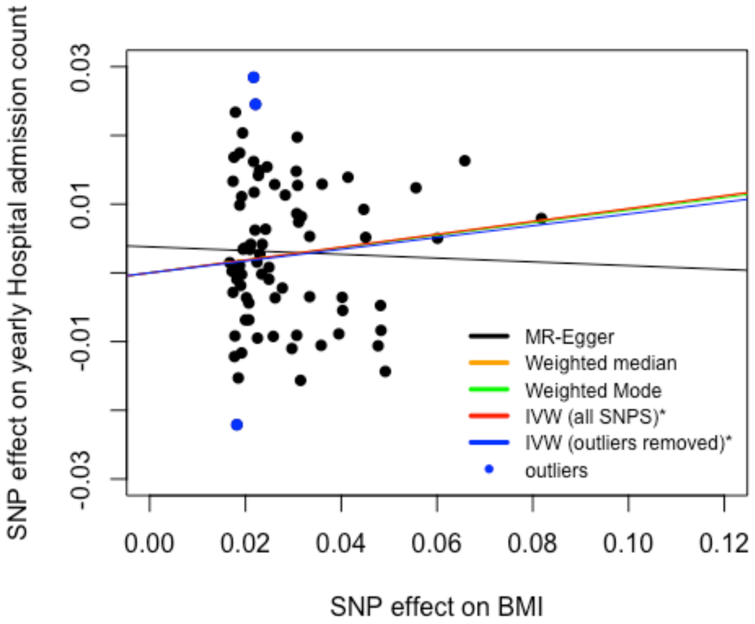

**Figure S2**. Plots for two-sample MR analysis of WHR effect (per SD, with $SD_{WHR}$=0.07) on yearly hospital admission rate in UK Biobank participants of White British ancestry. Effects are shown on the log(rate) scale. A) Cochran's Q is plotted against Rucker's Q for each SNP, as calculated in a leave-one-out analysis. Outliers have been visually identified and are labeled. Q-statistics for the model with the full sets of 34 SNPs (red) and with outliers removed (30 SNPs, blue) are shown. B) SNP effect on yearly hospital admission count is plotted against the SNP effect on WHR. Fitted lines are displayed for the random effects MR-Egger, penalized weighted median, weighted mode and random effects exact weight IVW models. The outliers identified in plot *A* are shown in blue. * denotes a significant slope.

**WHR**

**A**

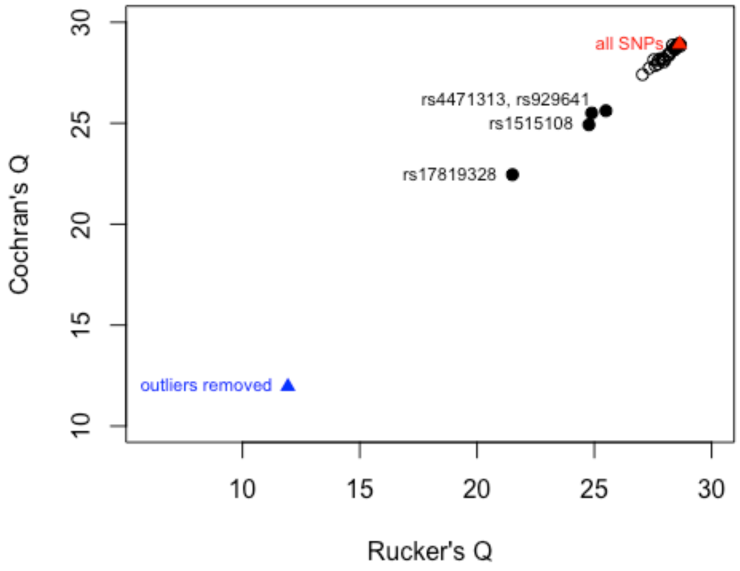


**B**

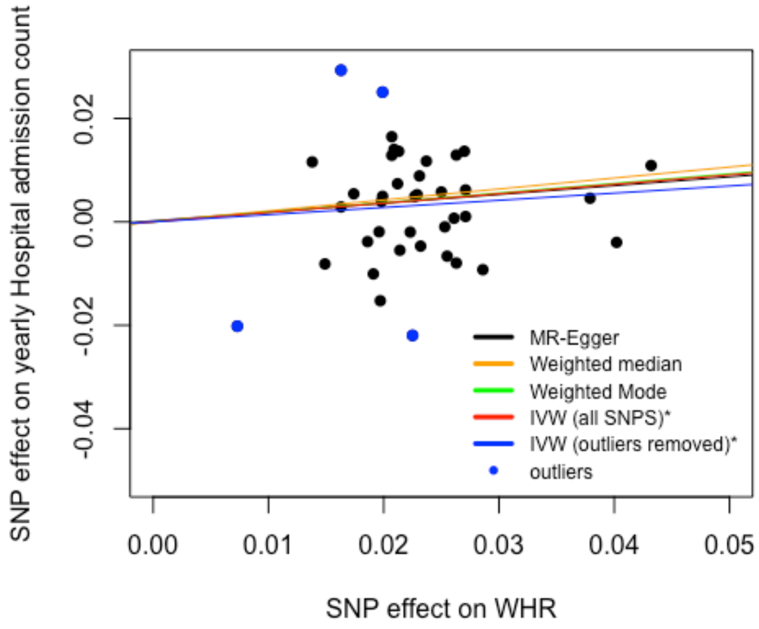

**Figure S3**. Plots for two-sample MR analysis of WHRadjBMI effect (per SD, with $SD_{WHR}$=0.07) on yearly hospital admission rate in UK Biobank participants of White British ancestry. Effects are shown on the log(rate) scale. A) Cochran's Q is plotted against Rucker's Q for each SNP, as calculated in a leave-one-out analysis. Outliers have been visually identified and are labeled. Q-statistics for the model with the full set of 45 SNPs (red) and with outliers removed (41 SNPs, blue) are shown. B) SNP effect on yearly hospital admission count is plotted against the SNP effect on WHRadjBMI. Fitted lines are displayed for the random effects MR-Egger, penalized weighted median, weighted mode and random effects exact weight IVW models. The outliers identified in plot *A* are shown in blue. * denotes a significant slope.

#### WHRadjBMI

**A**

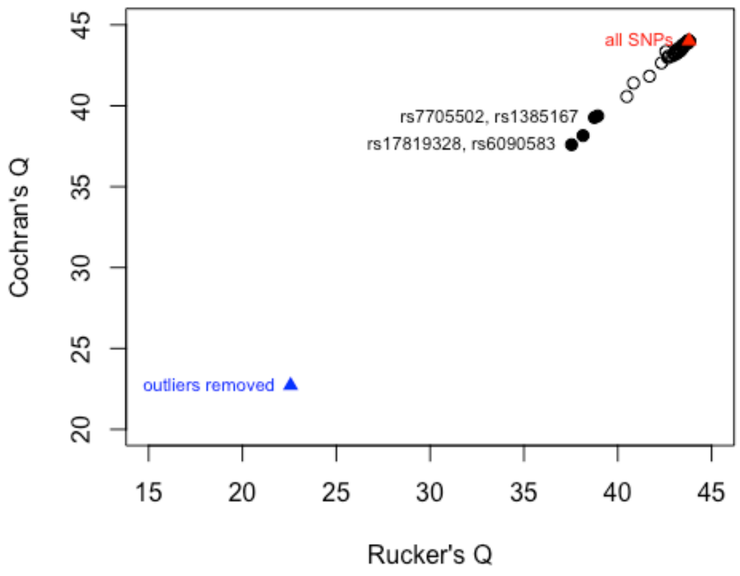


**B**

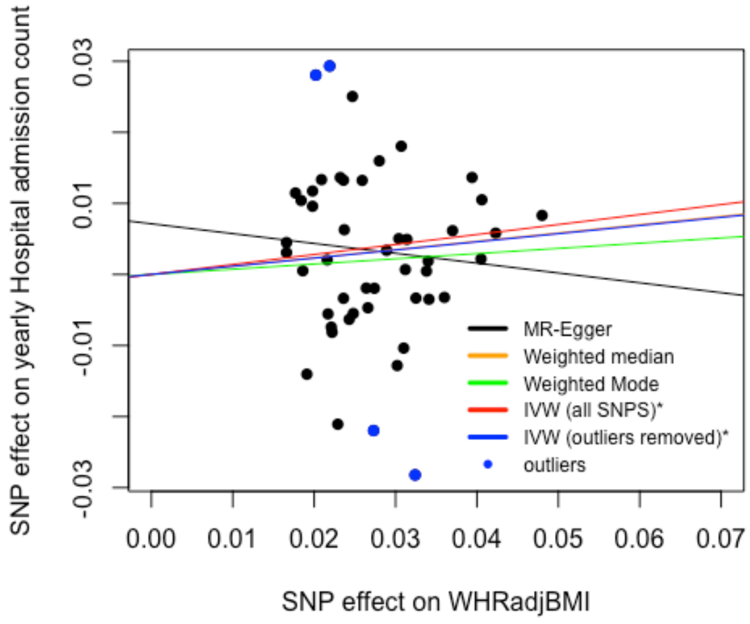
