## Supplementary material for "Mendelian Randomization analysis of the causal impact of body mass index and waist-hip ratio on rates of hospital admission": R code Appendix

### Code appendix

This document provides the necessary code for recreating the analyses reported in our paper. For simplicity, the univariable analyses have been restricted to those of a single exposure (BMI).

#### Univariable analyses

##### Observational analyses

data.red contains for every patient the baseline variable measurements and BMI and WHR measurements. Below the first 6 entries of the data are shown NTS add total.adm and person_years to data.red

```
##      n_eid total_adm person_years    sex       whr     bmi      age  townsend
## 9  1000049         1     6.327173   Male 0.9217391 34.5119 60.62971 -2.910100
## 10 1000065         1     5.861739   Male 0.8421053 25.6931 48.42710 -3.423470
## 20 1000082         2     6.899384 Female 0.8181818 32.5391 60.97741 -3.729590
## 28 1000134         3     4.709103 Female 0.7407407 26.6299 67.33470 -0.525358
## 31 1000142         4     5.552361   Male 0.9819820 33.8021 68.90349 -3.186940
## 38 1000178         1     5.645448 Female 0.8879311 33.4602 67.39767 -1.902140
##    days_exercise         alcochol_frequency
## 9              1     Special occasions only
## 10             3      Daily or almost daily
## 20             4       Once or twice a week
## 28             0      Daily or almost daily
## 31             0 One to three times a month
## 38             3       Once or twice a week
##                                              qualifications
## 9  Other professional qualifications eg: nursing, teaching
## 10                             O levels/GCSEs or equivalent
## 20                             O levels/GCSEs or equivalent
## 28                             College or University degree
## 31                             O levels/GCSEs or equivalent
## 38                             O levels/GCSEs or equivalent
##                               employment
## 9                                Retired
## 10 In paid employment or self-employed
## 20                               Retired
## 28                               Retired
## 31                               Retired
## 38                               Retired
```

```
## [1] 57.40228
```

```
## [1] 27.38488
```

```
##    Female      Male
## 0.5366446 0.4633554
```

5 of the baseline variables have missing values and are imputed in a ten-fold imputation. alcohol frequency, qualifications, employment and sex are treated as categorical variables, days exercise is treated as a ordinal one, while the townsend deprivation score is treated as a continuous variable (does not need to be explicitly specified in the function as continuous is the default). The ID variable (n_eid) is ignored in the imputation, as is the admission count (total_adm) and the years on study (person_years), while the observed values for BMI and WHR (and the other variables when measured) are allowed to influence the imputation process. Post imputation, the townsend deprivation variable is structured into quintiles. This is done after imputing, to allow the imputation process to be as accurate as possible. Additionally, the function mod.func restructures the factor levels of various variables with the purpose of defining logical reference categories.

```
# Imputation
#install.packages(amelia)
library(Amelia)
comp.dat <- amelia(data.red, m=10, noms=c("alcohol_frequency", "qualifications",
                                          "employment","sex"),
                                ord=c("days_exercise"),
                                idvars=c("n_eid", "total_adm", "person_years"))
colnames(data.red)
comp.dat <- comp.dat$imputations
comp.datMR <- comp.dat

# Restructuring each of the 10 imputed datasets
mod.func <- function(comp.dat, data.bmi){
  town <- comp.dat$townsend_deprivation
  qq <- as.numeric(quantile(town, probs = seq(0, 1, .2), na.rm=TRUE))
  new.town <- rep(NA, length(town))
  new.town[which(town>=qq[1] & town<=qq[2])] <- 1
  new.town[which(town>qq[2] & town<=qq[3])] <- 2
  new.town[which(town>qq[3] & town<=qq[4])] <- 3
  new.town[which(town>qq[4] & town<=qq[5])] <- 4
  new.town[which(town>qq[5] & town<=qq[6])] <- 5
  comp.dat$townsend_deprivation <- new.town
  comp.dat$townsend_deprivation <- as.factor(comp.dat$townsend_deprivation)
  comp.dat$alcohol_frequency <- relevel(comp.dat$alcohol_frequency,
                                        "Once or twice a week")
  comp.dat$employment <- relevel(comp.dat$employment,
                                 "In paid employment or self-employed")
  comp.dat$days_exercise <- as.factor(comp.dat$days_exercise)
  return(comp.dat)
}

comp.datMR <- lapply(comp.datMR, function(x) mod.func(x,data.bmi))
save(comp.datMR, file="comp.datMR")
```

A Poisson regression is performed on each of the 10 datasets, adjusting for baseline covariates with the log time on study (in years) as an offset. The estimates and standard errors are subsequently pooled using Rubin's rules.

```
# function that fits model
model.function.bmi <- function(comp.dat){
```

```r
  model.obj <- glm(total_adm ~ bmi_combined + sex + qualifications +
                   employment + townsend_deprivation +
                   days_exercise + alcohol_frequency + age +
                   offset(log(person_years)), family=quasipoisson,
                      data=comp.dat)
  print(Sys.time())
  return(model.obj)}

load(file="comp.datMR")

# fitting models
bmi.obs.models <- lapply(comp.datMR, function(x) model.function.bmi(x))

# pooling estimates
pooling.function <- function(model.list){

  m <- length(model.list)

  rowstab <- nrow(summary(model.list[[1]])$coefficients)
  # extracting coefficients and obtaining mean
  coefs <- lapply(model.list, coef)
  coefs <- data.frame(matrix(unlist(coefs), nrow=rowstab, byrow=FALSE),
                    stringsAsFactors=FALSE)
  mcoef <- apply(coefs, 1, mean)

  # calculating within, between and total variance according to Rubin's rule
  vars <- lapply(model.list, vcov)
  # below an alternative way to obtain the variance is given, depending on
  # R version the formermay misfire
  #vars <- lapply(model.list, function(x){
  #                              x <- x$var
  #                              return(x)
  #                                })
  within <- Reduce('+', vars)/m
  between <- lapply(coefs, function(x) (x-mcoef) %*% (t(x-mcoef)))
  between <- Reduce('+', between)/(m-1)
  tot <- within + ((m+1)/m) * between
  totSE <- as.numeric(sqrt(diag(tot)))

  # obtaining hazard ratios, confidence interval and P value
  Rate <- exp(mcoef)
  lower <- exp(mcoef-1.96*totSE)
  upper <- exp(mcoef+1.96*totSE)
  Zs <- mcoef/totSE
  Ps <- pnorm(abs(Zs),lower.tail=FALSE)*2

  # making table
  tab <- cbind(Rate, lower, upper,
             format.pval(Ps, eps=0.001, digits=3),
             mcoef, totSE)
  colnames(tab) <- c("Rate", "lower", "upper", "P value", "Coef", "SE")
  rownames(tab) <- rownames(as.data.frame(coef(model.list[[1]])))
```

```r
  return(tab)
}

bmi.obs.tab <- pooling.function(bmi.obs.models)
save(bmi.obs.tab, file="bmi.obs.tab")
load(file="bmi.obs.tab")


# obtain estimates from table with pooled estimates:
# point estimate, 95% CI upper limit, 95% CI lower limit, respectively
bmi.obs.c <- round(as.numeric(bmi.obs.tab[2,1]),3)
bmi.obs.c.u <- round(as.numeric(bmi.obs.tab[2,3]),3)
bmi.obs.c.l <- round(as.numeric(bmi.obs.tab[2,2]),3)

# obtain estimates on SD scale:
bmi.obs.cs <- round(exp(as.numeric(bmi.obs.tab[2,5])*sd(massive.bmi$bmi)),3)
bmi.obs.cs.u <- round(exp(as.numeric(bmi.obs.tab[2,5])*sd(massive.bmi$bmi) +
                        1.96*as.numeric(bmi.obs.tab[2,6])*sd(massive.bmi$bmi)),3)
bmi.obs.cs.l <- round(exp(as.numeric(bmi.obs.tab[2,5])*sd(massive.bmi$bmi) -
                        1.96*as.numeric(bmi.obs.tab[2,6])*sd(massive.bmi$bmi)),3)
```

In addition to adjusted estimates, estimates are also obtained without adjusting for baseline covariates.

```r
obs.pois <- glm(total_adm ~ bmi + offset(log(person_years)), family=quasipoisson,
                data=massive.bmi)
obs.pois.sc <- glm(total_adm ~ bmisc + offset(log(person_years)), family=quasipoisson,
                   data=massive.bmi)

# obtain uncorrected observational estimates on unit scale
bmi.obs <- round(exp(summary(obs.pois)$coefficients[2,1]),3)
bmi.obs.u <- round(exp(summary(obs.pois)$coefficients[2,1] +
                         1.96*summary(obs.pois)$coefficients[2,2]),3)
bmi.obs.l <- round(exp(summary(obs.pois)$coefficients[2,1] -
                         1.96*summary(obs.pois)$coefficients[2,2]),3)

# obtain uncorrected observational estimates on SD scale
bmi.obs.sc <- round(exp(summary(obs.pois.sc)$coefficients[2,1]),3)
bmi.obs.sc.u <- round(exp(summary(obs.pois.sc)$coefficients[2,1] +
                            1.96*summary(obs.pois.sc)$coefficients[2,2]),3)
bmi.obs.sc.l <- round(exp(summary(obs.pois.sc)$coefficients[2,1] -
                            1.96*summary(obs.pois.sc)$coefficients[2,2]),3)
```

#### One-sample MR analyses

For the one-sample MR analyses gene risk scores (GRS) are created. For each of the exposures individual level data for the relevant SNPs is available from UK Biobank. For BMI the SNP scores are contained in the dataframe data.bmi - an excerpt is shown below. Each column represents a genetic variant, each row a patient.

```
##  [1] "n_eid"        "total_adm"    "age.at"       "person_years" "n_eid_g"
##  [6] "bmi"          "whr"          "linkedid"     "rs657452_G"   "rs11583200_T"
## [11] "rs3101336_C"  "rs12566985_A" "rs12401738_A" "rs11165643_T" "rs17024393_C"
```

```
## [16] "rs543874_G"   "rs2820292_C"  "rs13021737_G" "rs10182181_G" "rs11126666_A"
## [21] "rs1016287_C"  "rs11688816_A" "rs2121279_T"  "rs1528435_T"  "rs7599312_A"
## [26] "rs6804842_G"  "rs2365389_T"  "rs3849570_A"  "rs13078960_G" "rs16851483_T"
## [31] "rs1516725_C"  "rs10938397_G" "rs17001654_G" "rs13107325_T" "rs11727676_C"
## [36] "rs2112347_G"  "rs205262_G"   "rs2033529_G"  "rs2207139_G"  "rs9400239_C"
## [41] "rs13191362_G" "rs1167827_G"  "rs2245368_T"  "rs17405819_C" "rs2033732_C"
## [46] "rs4740619_C"  "rs10968576_G" "rs6477694_T"  "rs1928295_C"  "rs10733682_G"
## [51] "rs7899106_G"  "rs17094222_C" "rs11191560_C" "rs7903146_T"  "rs4256980_G"
## [56] "rs11030104_G" "rs2176598_C"  "rs3817334_T"  "rs12286929_G" "rs7138803_A"
## [61] "rs11057405_A" "rs12429545_A" "rs10132280_A" "rs12885454_A" "rs11847697_T"
## [66] "rs7141420_T"  "rs3736485_G"  "rs16951275_C" "rs758747_T"   "rs12446632_A"
## [71] "rs2650492_A"  "rs3888190_A"  "rs9925964_G"  "rs1558902_A"  "rs1000940_G"
## [76] "rs12940622_A" "rs1808579_T"  "rs7243357_G"  "rs6567160_C"  "rs17724992_G"
## [81] "rs29941_G"    "rs2075650_G"  "rs2287019_T"  "rs3810291_A"  "GRS.bmi"
## [86] "sex"
```

```
##      n_eid rs657452_G rs11583200_T rs3101336_C rs12566985_A rs12401738_A
## 9  1000049          1            1           1            2      1.00000
## 10 1000065          1            1           1            1      0.00392
## 20 1000082          1            1           1            1      1.00000
## 28 1000134          0            0           2            2      0.00000
## 31 1000142          1            1           1            1      0.00000
## 38 1000178          0            0           2            1      1.00000
##    rs11165643_T rs17024393_C
## 9             1            0
## 10            1            0
## 20            2            0
## 28            1            0
## 31            2            0
## 38            0            0
```

Prior to calculating the GRS, the gene scores are recoded to ensure they represent the exposure increasing allele and are consistent with the gene-exposure coefficients retrieved from the GWAS meta-analyses (for BMI here the Locke 2016 study). These coefficients are contained in the object coefs.bmi. Below the GRS is generated. The GRS value for each patient is the average number of BMI-increasing alleles per individual.

```
data.w.bmi <- data.bmi[,9:84]

# the score for each genetic variant is multiplied by the coefficient.
for (i in 1:ncol(data.w.bmi)){
  data.w.bmi[,i] <- data.w.bmi[,i]*coefs.bmi[i]
}

# then the modified scores are summed for each patient.
GRS.bmi <- apply(data.w.bmi,1,sum)

# then the modified scores are rescaled by dividing them by the sum of all the SNP effect
# sizes and multiplying with the total number of SNPs (76).
# this will give the average number of BMI increasing alleles per individual.
GRS.bmi <- (GRS.bmi/sum(coefs.bmi))*length(coefs.bmi)

data.bmi <- cbind(data.bmi, GRS.bmi)
```

Having obtained the GRS, the one-sample MR analyses are preformed, on the unit and SD scale, with adjustment for sex, age and the first 40 genetic PCAs. IV estimates are obtained using the Wald ratio approach, with the first stage (gene-exposure) linear, and the second stage (gene-outcome) a Poisson model with the log time on study (in years) as an offset. As the Poisson model is linear on the log scale, the ratio of the log(gene-outcome association) over the gene-exposure association can be taken, and then exponentiated to yield the IV rate estimate.

```r
load(file="massive.bmi")
massive.bmi$bmisc <- scale(massive.bmi$bmi)
save(massive.bmi, file="massive.bmi")

# vector with all PCA names
# (of the form "X.12.1725 + X5.39163 + X.1.28103 + ...+ X.3.55047 + ")
formbmi <- paste(paste(colnames(massive.bmi)[87:126], " + ", sep=""), collapse="")

# formula for the regression of outcome hospital admission count on the BMI GRS,
# 40 PCAs, age and sex, with the log years on study as offset
big.formula <- paste(paste(paste(paste("total_adm ~ ", "GRS.bmi",sep = ""), " + ", sep=""),
                  formbmi, sep=""), "age.at + sex + offset(log(person_years))", sep="")

# formula for the regression of exposure BMI on the BMI GRS, 40 PCAs, age and sex
big.formula.exp <- paste(paste(paste(paste("bmi ~ ", "GRS.bmi",sep = ""), " + ", sep=""),
                    formbmi, sep=""), "age.at + sex", sep="")

# formula for the regression of exposure BMI on the BMI GRS, 40 PCAs, age and sex,
# with BMI on the SD scale
big.formula.expsc <- paste(paste(paste(paste("bmisc ~ ", "GRS.bmi",sep = ""), " + ", sep=""),
                                 formbmi, sep=""), "age.at + sex", sep="")

# regression outcome on GRS, adjusting for age, sex and first 40 genetic PCAs
out.pois <- glm(big.formula, family=quasipoisson, data=massive.bmi)

# regression exposure on unit scale on GRS, adjusting for age, sex and first 40 genetic
# PCAs
exp.lin <- lm(big.formula.exp, data=massive.bmi)

# regression exposure on SD scale on GRS, adjusting for age, sex and first 40 genetic PCAs
exp.lin.sc <- lm(big.formula.expsc, data=massive.bmi)


# unit scale coefficients and standard errors
Beta_num <- summary(out.pois)$coefficients[2,1]
SE_num <- summary(out.pois)$coefficients[2,2]
Beta_den <- summary(exp.lin)$coefficients[2,1]
SE_den <- summary(exp.lin)$coefficients[2,2]

# the SE was estimated using this taylor series expansion based formula
SE_IV <- sqrt((SE_num^2/Beta_den^2) + (Beta_num^2/Beta_den^4)*SE_den^2 -
                2*(Beta_num/Beta_den^3)*0)

# point estimate and upper and lower 95% confidence interval values for BMI on the
#unit scale
bmi.IV <- round(exp(Beta_num/Beta_den),3)
bmi.IV.l <- round(exp((Beta_num/Beta_den)-SE_IV*1.96),3)
bmi.IV.u <- round(exp((Beta_num/Beta_den)+SE_IV*1.96),3)
```

```r
# scaled coefficients and standard errors
Beta_num_sc <- summary(out.pois)$coefficients[2,1]
SE_num_sc <- summary(out.pois)$coefficients[2,2]
Beta_den_sc <- summary(exp.lin.sc)$coefficients[2,1]
SE_den_sc <- summary(exp.lin.sc)$coefficients[2,2]

# the SE was estimated using this taylor series expansion based formula
SE_IV_sc <- sqrt((SE_num_sc^2/Beta_den_sc^2) + (Beta_num_sc^2/Beta_den_sc^4)*
                   SE_den_sc^2 -2*(Beta_num_sc/Beta_den_sc^3)*0)

# point estimate and upper and lower 95% confidence interval values for BMI on the
#SD scale
bmi.sc.IV <- round(exp(Beta_num_sc/Beta_den_sc),3)
bmi.sc.IV.u <- round(exp((Beta_num_sc/Beta_den_sc)+SE_IV_sc*1.96),3)
bmi.sc.IV.l <- round(exp((Beta_num_sc/Beta_den_sc)-SE_IV_sc*1.96),3)
```

This procedure is repeated for the exposure WHR, using both WHR and WHRadjBMI SNPs. With the latter set of SNPs an additional exposure is considered- the residuals from a linear regression of WHR on BMI. Below an example of this regression is shown for WHR on the SD scale (with whrsc the scaled WHR).

```r
first.reg <- lm(whrsc ~bmi, data=massive.bmi.f)
resids.sc <- residuals(first.reg)
```

#### Two-sample summary MR analyses

```r
library(TwoSampleMR)
```

```
## TwoSampleMR version 0.5.5
## [>] New: Option to use non-European LD reference panels for clumping etc
## [>] Some studies temporarily quarantined to verify effect allele
## [>] See news(package='TwoSampleMR') and https://gwas.mrcieu.ac.uk for further details
```

```r
library(MRInstruments)
library(RadialMR)
```

Objects twosampBMI, twosampWHR and twosampadjWHR contain the exposure and outcome coefficients and standard errors for BMI, WHR and WHRadjBMI, respectively, and are formatted as TwoSampleMR harmonise_exposure_outcome objects. Effect alleles have been defined so that they reflect exposure-increasing coefficients. The WHR and WHRadjBMI coefficients and standard errors were obtained from the Shungin et al. (2015) GWAS meta-analysis, the BMI coefficients from the Locke et al. (2016) GWAS meta-analysis. The outcome coefficients and standard errors were calculated from the UK Biobank data, using individual level genetic data and adjusting for sex, age and the first 40 genetic principle components. The default scale for the exposure coefficients is SD for all three exposures. To obtain IV estimates on a different scale, one can either rescale the relevant exposure-gene association coefficients, or rescale the IV estimate itself on the log scale.

```r
load(file="twosampBMI")
load(file="twosampWHR")
load(file="twosampwhr.adj")
```

```r
head(twosampBMI)
```

```
##   beta.outcome se.outcome id.exposure id.outcome        SNP beta.exposure
## 1  0.014167799 0.01059486         BMI      COUNT   rs657452        0.0227
## 2 -0.012180115 0.01062840         BMI      COUNT rs11583200        0.0177
## 3  0.005306914 0.01053646         BMI      COUNT  rs3101336        0.0334
## 4  0.006347829 0.01041519         BMI      COUNT rs12566985        0.0242
## 5  0.004163216 0.01064471         BMI      COUNT rs12401738        0.0211
## 6  0.011723487 0.01051890         BMI      COUNT rs11165643        0.0218
##   se.exposure mr_keep exposure outcome  NA                  NA EA OA   EAF
## 1      0.0031    TRUE        1       2 BMI Hospital admissions  A  G 0.394
## 2      0.0031    TRUE        1       2 BMI Hospital admissions  C  T 0.396
## 3      0.0031    TRUE        1       2 BMI Hospital admissions  C  T 0.613
## 4      0.0031    TRUE        1       2 BMI Hospital admissions  G  A 0.446
## 5      0.0033    TRUE        1       2 BMI Hospital admissions  A  G 0.352
## 6      0.0031    TRUE        1       2 BMI Hospital admissions  T  C 0.583
```

The SNP data is clumped, using an LD threshold of $R2 < 0.001$. The clumped object can be fed directly to the TwoSampleMR mr() function, here used to obtain the random effects MR-Egger and weighted mode estimates. The ivw_radial() function from the RadialMR package is used to obtain the exact weights random effects IVW estimate. Prior to this, the clumped data must be reformatted using the format_radial function. The mr_penalised_weighted_median() function is used to obtain the penalized weighted median estimate. Here an example is shown for BMI estimates obtained on the SD scale (the default, the Locke 2016 study gives exposure coefficients on the SD scale). To obtain estimates on the unit scale, adjustments are made to the twosampBMI dataset (e.g. prior to MR analysis, as the estimates from the MR estimators cannot be transformed for such a purpose)

```r
devtools::install_github("MRCIEU/TwoSampleMR")
devtools::install_github("WSpiller/RadialMR")
library(TwoSampleMR)
library(MRInstruments)
library(RadialMR)

# clump data
exp.BMI.clump <- clump_data(twosampBMI[,c(2,3,5,6,7,9)])
temp.BMIad <- twosampBMI[which(twosampBMI$SNP %in% exp.BMI.clump$SNP),]
exp.BMI.clump <- cbind(exp.BMI.clump, temp.BMIad[,c(1,4,8,10)])

nrow(twosampBMI)
SNP.id <- twosampBMI$SNP[which(twosampBMI$SNP %in% c("rs7243357", "rs205262", "rs12940622")) ]
#SNP.id[3] <- twosampBMI$SNP[27]
exp.BMI.clump <- exp.BMI.clump[-which(exp.BMI.clump$SNP %in% SNP.id),]
nrow(exp.BMI.clump)

# format data for RadialMR package
exp.BMI <- format_radial(exp.BMI.clump$beta.exposure,
                         exp.BMI.clump$beta.outcome,
                         exp.BMI.clump$se.exposure,
                         exp.BMI.clump$se.outcome, exp.BMI.clump$SNP)

save(exp.BMI.clump, file="exp.BMI.clump")
save(exp.BMI, file="exp.BMI")
```

```
# obtain MR-Egger, weighted mode, penalized weighted median and exact weights
# random effects IVW estimates with 95% confidence intervals

# MR-Egger intercept and slope and weighted mode estimates
res.BMI <- mr(exp.BMI.clump)
intercept.Egg.BMI <- mr_pleiotropy_test(exp.BMI.clump)
BMI.egg.int <- cbind(exp(intercept.Egg.BMI$egger_intercept),
      exp(intercept.Egg.BMI$egger_intercept- 1.96* intercept.Egg.BMI$se),
      exp(intercept.Egg.BMI$egger_intercept+ 1.96* intercept.Egg.BMI$se))


rel.res.BMI <- rbind(BMI.egg.int,
            cbind(exp(res.BMI$b), exp(res.BMI$b - 1.96*res.BMI$se),
                  exp(res.BMI$b + 1.96*res.BMI$se))[c(1,5),])

# penalized weighted median and IVW estimatse
BMI.pen.med <- mr_penalised_weighted_median(exp.BMI.clump$beta.exposure,
                                            exp.BMI.clump$beta.outcome,
                                            exp.BMI.clump$se.exposure,
                                            exp.BMI.clump$se.outcome,
                                      parameters=default_parameters())
BMI.pen.med.e <- cbind(exp(BMI.pen.med$b),
                       exp(BMI.pen.med$b - 1.96*BMI.pen.med$se),
                       exp(BMI.pen.med$b + 1.96*BMI.pen.med$se))
BMI.IVW <- ivw_radial(exp.BMI, weights=3)
BMI.IVW.e <- cbind(exp(BMI.IVW$coef$Estimate[4]),
                   exp(BMI.IVW$coef$Estimate[4]-1.96*BMI.IVW$coef$Std.Error[4]),
                   exp(BMI.IVW$coef$Estimate[4]+1.96*BMI.IVW$coef$Std.Error[4]))


# obtaining output
BMI.SD.output <- rbind(rel.res.BMI[c(1:2),], BMI.IVW.e, BMI.pen.med.e,
                       rel.res.BMI[3,])
BMI.SD.output <- round(BMI.SD.output, 3)
BMI.SD.output.obj <- paste(paste(paste(paste(BMI.SD.output[,1], "(", sep=" "),
              BMI.SD.output[,2], sep=""),BMI.SD.output[,3], sep="-"), ")",
              sep="")
```

Heterogeneity is evaluated using the IVW-derived Cochran's Q statistic and the MR-Egger derived Rucker's Q statistic. Both are obtained using the ivw_radial() and ivw_egger() functions from the RadialMR package, which, in addition to the coefficients, generate Cochran's Q and Rucker's Q, respectively. Both Q statistics are obtained for each SNP in a leave-one-out sensitivity analysis.

```
# Start with data frame with following columns: SNP beta.exposure  beta.outcome
# se.exposure  se.outcome. Let's call this data frame exp.BMI
# Then create an indicator vector in matrix form (so 1 long column )
ind3 <- as.matrix(seq(1,nrow(exp.BMI)))

# this function takes an index 'ind' and removes the corresponding SNP,
# and returns the Q statistics for IVW and Egger after fitting the models to the
# remaining SNPS
rad.func3 <- function(ind){
  dat <- exp.BMI[-ind,]
```

```
  rad.whr.ivw <- ivw_radial(dat, weights=3)
  rad.whr.egg <- egger_radial(dat,weights=3)
  Q <- rad.whr.ivw$qstatistic
  Q2 <- rad.whr.egg$qstatistic
  print(Sys.time())
  return(c(Q, Q2))
}

# applying the function using apply
one.out.bmi <- apply(ind3,1,rad.func3)
save(one.out.bmi, file="one.out.bmi22")
load(file="one.out.bmi22")

# restructuring output
outss.bm <- as.data.frame(t(one.out.bmi))
colnames(outss.bm) <- c("y", "x") # y is IVW, x=Egger

# adding in SNP names
final.out <- cbind(exp.BMI$SNP, outss.bm)
colnames(final.out) <- c("SNP", "IVW", "Egger")

# scatterplot: identifying outliers visually
plot(final.out[,2]~final.out[,3], ylab="Cochran's Q", xlab="Rucker's Q")

# order according to IVW
final.out[order(final.out[,2]),]

# order according to Egger
final.out[order(final.out[,3]),]

# Fitting IVW and Egger models to full data and obtaining Q-statistics
rad.bmi.ivw <- ivw_radial(exp.BMI)
rad.bmi.egg <- egger_radial(exp.BMI)
rad.bmi.ivw$qstatistic
rad.bmi.egg$qstatistic

# remove outliers
new.dat.bmi <- exp.BMI[-c(59, 25, 61),] #34
rad.bmi.ivw.new <- ivw_radial(new.dat.bmi, weights=3)
rad.bmi.egg.new <- egger_radial(new.dat.bmi, weights=3)
final.out$SNP <- as.character(final.out$SNP)
final.outF <- final.out

# create plot
plot(final.outF$IVW~final.outF$Egger, ylab="Cochran's Q", xlab="Rucker's Q",
     ylim=c(30,55), xlim=c(28,55))
points(x=45.43864,y=46.74076, pch=16) #18
points(x=47.25262,y=47.67067, pch=16) #29
points(x=47.82142,y=48.32312, pch=16) #38
points(x=rad.bmi.egg$qstatistic,y=rad.bmi.ivw$qstatistic, pch=17, col="red")
points(x=rad.bmi.egg.new$qstatistic,y=rad.bmi.ivw.new$qstatistic, pch=17,
       col="blue")
```

```r
final.new <- rbind(final.out,c("total",rad.bmi.ivw$qstatistic,
                               rad.bmi.egg$qstatistic),
                         c("total",rad.bmi.ivw.new$qstatistic,
                           rad.bmi.egg.new$qstatistic))

final.new <- as.data.frame(final.new)
final.new$IVW <- as.numeric(final.new$IVW)
final.new$Egger <- as.numeric(final.new$Egger)

with(final.new[c(59,25,65,66),], text(IVW~Egger,
     labels = c("rs12940622","rs7243357, rs205262","all SNPs",
    "outliers removed"), pos = c(2,2,2,2,2), col=c(rep("black",2),
                                    "red", "blue"), cex=0.7))


# visual illustration estimates (SNP effect on outcome plotted against
# SNP effect on BMI)
plot(exp.BMI.clump$beta.outcome~exp.BMI.clump$beta.exposure,
     ylab="SNP effect on yearly Hospital admission count",
     xlab="SNP effect on BMI",
     pch=16, xlim=c(0,0.13), ylim=c(-0.05,0.03))
abline(a=0.003785982, b=-0.02763678, col="black")
abline(a=0, b= 0.09352181, col="orange")
abline(a=0, b=0.09112850, col="green")
abline(a=0, b=0.09342596 , col="red")
abline(a=0, b=0.08570039, col="blue")

index.bmi <- seq(1,nrow(exp.BMI.clump))[-which(exp.BMI.clump$SNP %in%
                                              new.dat.bmi$SNP)]
points(exp.BMI.clump$beta.exposure[index.bmi],
       exp.BMI.clump$beta.outcome[index.bmi], col="blue", pch=16)

legend(x=0.07, y=-0.01, c("MR-Egger", "Weighted median", "Weighted Mode",
                "IVW (all SNPS)*", "IVW (outliers removed)*", "outliers"),
                  col=c("black", "orange", "green", "red", "blue", "blue"),
                     lty=c(1,1,1,1,1,NA),lwd=c(3,3,3,3,3,NA),
                      pch=c(NA,NA,NA,NA,NA,16), cex=0.75, bty="n")


# Remove SNPs from data, create objects red.exp.BMI.clump and
# red.exp.BMI and repeat analysis in previous code chunk
# (sensitivity analysis)
rem.SNP.BMI <- c("rs12940622","rs7243357", "rs205262")

load(file="exp.BMI.clump")
load(file="exp.BMI")

exp.BMI.clump$beta.exposure <- exp.BMI.clump$beta.exposure*4.6
exp.BMI.clump$se.exposure <- exp.BMI.clump$se.exposure*4.6
exp.BMI <- format_radial(exp.BMI.clump$beta.exposure,
                         exp.BMI.clump$beta.outcome,
                         exp.BMI.clump$se.exposure,
                         exp.BMI.clump$se.outcome, exp.BMI.clump$SNP)
```

```
red.exp.BMI.clump <- exp.BMI.clump[-
                          which(exp.BMI.clump$SNP %in% rem.SNP.BMI),]
red.exp.BMI <- exp.BMI[-which(exp.BMI$SNP %in% rem.SNP.BMI),]

# apply relevant MR methods
```

### Multivariable analyses

#### Observational Multivariable MR analyses

Observational multivariable estimates are obtained by a regression of the hospital admission count outcome on BMI and WHR jointly, adjusting for various baseline covariates and using the log time on observation (in years) as an offset. Adjusted estimates are obtained in a 10-fold multiple imputation approach (as for the univariable estimates). All estimates are obtained on the SD scale and the unit and 0.1 unit scale for BMI and WHR, respectively.

```
load(file="comp.datMR")

# model fitting function
model.function.mult <- function(comp.dat){
  model.obj <- glm(total_adm ~ bmi_combined + whr + sex + qualifications +
                     employment + townsend_deprivation + days_exercise +
                     alcohol_frequency + age + offset(log(person_years)),
                   family=quasipoisson, data=comp.dat)
  print(Sys.time())
  return(model.obj)}

# fitting models
mult.obs.models <- lapply(comp.datMR, function(x) model.function.mult(x))

# pooling function
pooling.function <- function(model.list){

  m <- length(model.list)

  rowstab <- nrow(summary(model.list[[1]])$coefficients)
  # extracting coefficients and obtaining mean
  coefs <- lapply(model.list, coef)
  coefs <- data.frame(matrix(unlist(coefs), nrow=rowstab, byrow=FALSE),
                      stringsAsFactors=FALSE)
  mcoef <- apply(coefs, 1, mean)

  # calculating within, between and total variance according to Rubin's rule
  vars <- lapply(model.list, vcov)
  within <- Reduce('+', vars)/m
  between <- lapply(coefs, function(x) (x-mcoef) %*% (t(x-mcoef)))
  between <- Reduce('+', between)/(m-1)
  tot <- within + ((m+1)/m) * between
  totSE <- as.numeric(sqrt(diag(tot)))

  # obtaining hazard ratios, confidence interval and P value
```

```
  Rate <- exp(mcoef)
  lower <- exp(mcoef-1.96*totSE)
  upper <- exp(mcoef+1.96*totSE)
  Zs <- mcoef/totSE
  Ps <- pnorm(abs(Zs),lower.tail=FALSE)*2

  # making table
  tab <- cbind(Rate, lower, upper,
               format.pval(Ps, eps=0.001, digits=3),
               mcoef, totSE)
  colnames(tab) <- c("Rate", "lower", "upper", "P value", "Coef", "SE")
  rownames(tab) <- rownames(as.data.frame(coef(model.list[[1]])))

  return(tab)
}

# pooled estimates
mult.obs.tab <- pooling.function(mult.obs.models)


# obtain adjusted estimates
# for BMI on unit scale
mult.obs.bmi.c <- round(as.numeric(mult.obs.tab[2,1]),3)
mult.obs.bmi.c.u <- round(as.numeric(mult.obs.tab[2,3]),3)
mult.obs.bmi.c.l <- round(as.numeric(mult.obs.tab[2,2]),3)

# for BMI on SD scale
mult.obs.bmi.cs <- round(exp(as.numeric(mult.obs.tab[2,5])*sd(massive.bmi$bmi)),3)
mult.obs.bmi.cs.u <- round(exp(as.numeric(mult.obs.tab[2,5])*sd(massive.bmi$bmi) +
                      1.96*as.numeric(mult.obs.tab[2,6])*sd(massive.bmi$bmi)),3)
mult.obs.bmi.cs.l <- round(exp(as.numeric(mult.obs.tab[2,5])*sd(massive.bmi$bmi) -
                      1.96*as.numeric(mult.obs.tab[2,6])*sd(massive.bmi$bmi)),3)

# for WHR on 0.1 unit scale
mult.obs.whr.c <- round(exp(as.numeric(mult.obs.tab[3,5])*0.1),3)
mult.obs.whr.c.u <- round(exp(as.numeric(mult.obs.tab[3,5])*0.1 +
                    1.96*as.numeric(mult.obs.tab[3,6])*0.1),3)
mult.obs.whr.c.l <- round(exp(as.numeric(mult.obs.tab[3,5])*0.1 -
                    1.96*as.numeric(mult.obs.tab[3,6])*0.1),3)

# for WHR on SD scale
mult.obs.whr.sc <- round(exp(as.numeric(mult.obs.tab[3,5])*sd(massive.bmi$whr)),3)
mult.obs.whr.sc.u <- round(exp(as.numeric(mult.obs.tab[3,5])*sd(massive.bmi$whr) +
                      1.96*as.numeric(mult.obs.tab[3,6])*sd(massive.bmi$whr)),3)
mult.obs.whr.sc.l <- round(exp(as.numeric(mult.obs.tab[3,5])*sd(massive.bmi$whr) -
                      1.96*as.numeric(mult.obs.tab[3,6])*sd(massive.bmi$whr)),3)


# obtaining unadjusted  estimates
massive.bmi$whrssc <- scale(massive.bmi$whr)
massive.bmi$whr01 <- (massive.bmi$whr)*10
obs.pois.mult <- glm(total_adm ~ bmi + whr01 + offset(log(person_years)),
                      family=quasipoisson, data=massive.bmi)
```

```r
obs.pois.mult.sc <- glm(total_adm ~ bmisc + whrssc + offset(log(person_years)),
                        family=quasipoisson, data=massive.bmi)

bmi.obs.mult.sc <- round(exp(summary(obs.pois.mult.sc)$coefficients[2,1]),3)
bmi.obs.mult.sc.u <- round(exp(summary(obs.pois.mult.sc)$coefficients[2,1] +
                             1.96*summary(obs.pois.mult.sc)$coefficients[2,2]),3)
bmi.obs.mult.sc.l <- round(exp(summary(obs.pois.mult.sc)$coefficients[2,1] -
                             1.96*summary(obs.pois.mult.sc)$coefficients[2,2]),3)

bmi.obs.mult <- round(exp(summary(obs.pois.mult)$coefficients[2,1]),3)
bmi.obs.mult.u <- round(exp(summary(obs.pois.mult)$coefficients[2,1] +
                         1.96*summary(obs.pois.mult)$coefficients[2,2]),3)
bmi.obs.mult.l <- round(exp(summary(obs.pois.mult)$coefficients[2,1] -
                         1.96*summary(obs.pois.mult)$coefficients[2,2]),3)

whr.obs.mult.sc <- round(exp(summary(obs.pois.mult.sc)$coefficients[3,1]),3)
whr.obs.mult.sc.u <- round(exp(summary(obs.pois.mult.sc)$coefficients[3,1] +
                         1.96*summary(obs.pois.mult.sc)$coefficients[3,2]),3)
whr.obs.mult.sc.l <- round(exp(summary(obs.pois.mult.sc)$coefficients[3,1] -
                         1.96*summary(obs.pois.mult.sc)$coefficients[3,2]),3)

whr.obs.mult <- round(exp(summary(obs.pois.mult)$coefficients[3,1]),3)
whr.obs.mult.u <- round(exp(summary(obs.pois.mult)$coefficients[3,1] +
                         1.96*summary(obs.pois.mult)$coefficients[3,2]),3)
whr.obs.mult.l <- round(exp(summary(obs.pois.mult)$coefficients[3,1] -
                         1.96*summary(obs.pois.mult)$coefficients[3,2]),3)
```

#### One-sample multivariable MR analysis

Multivariable MR in the one sample framework is performed by regressing each exposure separately on the joint set of genes (here regression BMI and WHR separately on the full set of combined BMI and WHR genes), obtaining the fitted values from each regression and then regressing the outcome (here hospital admission count) on the fitted values with the log time on study (years) as an offset. The analysis is performed with adjustment for age, sex and the first 40 genetic PCAs in both stages, with estimates obtained on the SD scale and on the unit and 0.1 unit scale for BMI and WHR, respectively. The dataset is called 'massive.mult' here, and contains the scores for the BMI and WHR genetic variants, the 40 genetic PCAs, the hospital admission count, the years on observation, BMI and WHR measurements and additional variables sex and age on entry. Variables are given in columns, with a row for each individual. Standard errors are obtained via bootstrap, by sampling 10000 datasets with replacement, calculating the IV estimates for BMI and WHR on each, and taking the SD for each of the exposures across the 10000 bootstraps.

```r
# loading data
load(file="massive.bmi")
load(file="massive.whr")

massive.mult <- cbind(massive.bmi, massive.whr)


# Creating formulas for first stage regression: this is done by selecting the
# relevant column names, alternatively the names can be specified manually, but
# this is more time consuming
# selecting gene names for bmi and whr
names.bmi <- colnames(massive.bmi)[9:84]
```

```r
names.whr <- colnames(massive.whr)[9:47]

# removing doubles
names.bmi <- names.bmi[-which(names.bmi %in% names.whr)]

# creating strings of form "gen1 + gene2 + ... + gene 3 +)
names.bmi <- paste(names.bmi, " + ", sep="", collapse="")
names.whr <- paste(names.whr, " + ", sep="", collapse="")

# selecting first 40 genetic PCA names
pcas <- paste(paste(colnames(massive.bmi)[87:126], " + ", sep=""), collapse="")

# pooling all to obtain string with genes, PCAs, age and sex
names.all <- paste(names.bmi, names.whr, sep="", collapse="")
for.form <- paste(names.all, pcas, sep="", collapse="")
for.form <- paste(for.form, "age.at + sex", sep="", collapse="")

# formula for regression BMI on the unit scale on all genes and 40 PCAs, age
# and sex
for.form.bmi <- paste("bmi ~ ", for.form, sep="", collapse="")

# formula for regression WHR on the 0.1 unit scale on all genes and 40 PCAs, age
# and sex
for.form.whr <- paste("whr01 ~ ", for.form, sep="", collapse="")


# First stage adjusted regressions
# first stage regressions on BMI unit scale and WHR 0.1 unit scale, adjusted,
# obtaining fitted values
lin.reg.mult <- lm(for.form.bmi, data=massive.mult)
lin.reg.mult.whr <- lm(for.form.whr, data=massive.mult)
fit.bmi <- fitted.values(lin.reg.mult)
fit.whr <- fitted.values(lin.reg.mult.whr)


# adding fitted values to dataframe, to be used in second stage regression
massive.mult$fit.bmi <- fit.bmi
massive.mult$fit.whr <- fit.whr

# creating formulas for second stage regression
# formulas  for regression outcome on fitted values BMI and WHR, adjusted for first
# 40 genetic PCAs, age and sex on unit scale
out.form <- paste("fit.bmi + fit.whr + ",
              paste(pcas, "age.at + sex + offset(log(person_years))", sep="",
                  collapse=""), sep="", collapse="")
out.form <- paste("total_adm ~ ", out.form)


# obtaining point estimates BMI and WHR on the unit scale (adjusted)
out.mult <- glm(out.form, family=quasipoisson, data=massive.mult)
BMIstat <- summary(out.mult)$coefficients[2,1:2]
WHRstat <- summary(out.mult)$coefficients[3,1:2]
STAT <- c(BMIstat,WHRstat)
```

```r
# performing bootstrap to obtain standard errors for confidence intervals

# loading data
load(file="massive.mult")

# creating list of 10000 with indexes for bootstrap samples
massive.index.list.a <- list()
for (i in 1:10000){
  index <- seq(1,nrow(massive.mult))
  dat.index <- sample(index, length(index), replace=TRUE)
  massive.index.list.a[[i]] <- dat.index
}

# bootstrap function. applies multivariable MR model to each bootstrapped dataset,
# taking as argument the bootstrap sample index. note that for.form.bmi, for.form.whr
# and out.form are the long formulas with genes, PCAs, sex and age that have been
# previously defined
boot.func.mult <- function(dat.index){
  dat <- massive.mult[dat.index,]

  lin.reg.mult <- lm(for.form.bmi, data=dat)
  lin.reg.mult.whr <- lm(for.form.whr, data=dat)
  fit.bmi <- fitted.values(lin.reg.mult)
  fit.whr <- fitted.values(lin.reg.mult.whr)

  dat$fit.bmi <- fit.bmi
  dat$fit.whr <- fit.whr

  out.mult <- glm(out.form, family=quasipoisson, data=massive.mult)
  BMIstat <- summary(out.mult)$coefficients[2,1:2]
  WHRstat <- summary(out.mult)$coefficients[3,1:2]
  STAT <- c(BMIstat,WHRstat)
  print(Sys.time())
  return(STAT)

}

# performing bootstrap (note this is computationally very intensive, recommend
# applying function to several smaller lists (rather than one list of 10.000)
# and/or saving partial objects during the function run
bootstraplist <- lapply(massive.index.list.a, boot.func.mult)

# obtaining bootstrap result for adjusted estimates
bootstraps <- do.call("rbind", bootstraplist)

# SD BMI
bmi.mult.sd <- sd(bootstraps[,1])

# point estimate and confidence interval BMI on unit scale
bmi.mult <- round(exp(STAT[1]),3)
bmi.mult.u <- round(exp(STAT[1] + bmi.mult.sd*1.96),3)
bmi.mult.l <- round(exp(STAT[1] - bmi.mult.sd*1.96),3)
```

```r
# point estimate and confidence interval BMI on SD scale
bmi.mult.sc <- round(exp(STAT[1]*sd(massive.mult$bmi)),3)
bmi.mult.sc.u <- round(exp(STAT[1]*sd(massive.mult$bmi) +
                             bmi.mult.sd * 1.96 * sd(massive.mult$bmi)),3)
bmi.mult.sc.l <- round(exp(STAT[1]*sd(massive.mult$bmi) -
                             bmi.mult.sd * 1.96 * sd(massive.mult$bmi)),3)


# SD WHR
whr.mult.sd <- sd(bootstraps[,3])

# point estimate and confidence interval WHR on 0.1 unit scale
whr01.mult <- round(exp(STAT[3] *0.10),3)
whr01.mult.u <- round(exp(STAT[3] *0.10 + whr.mult.sd*0.1*1.96),3)
whr01.mult.l <- round(exp(STAT[3] *0.10 - whr.mult.sd*0.1*1.96),3)

# point estimate and confidence interval WHR on SD scale
whr01.mult.sc <- round(exp(STAT[3] * sd(massive.mult$whr)),3)
whr01.mult.sc.u <- round(exp(STAT[3] * sd(massive.mult$whr) +
                               whr.mult.sd * 1.96 * sd(massive.mult$whr)),3)
whr01.mult.sc.l <- round(exp(STAT[3] * sd(massive.mult$whr) -
                               whr.mult.sd * 1.96 * sd(massive.mult$whr)),3)



# obtaining conditional F-statistics

# Conditional F statistic WHR

# 1) BMI is regressed on full set of genetic instruments + control
# variables, predicted values are obtained
test1a <- paste("bmi ~ ", for.form, sep="", collapse="")
obj1a <- lm(test1a, data=massive.mult)
preds2 <- fitted.values(obj1a)
massive.mult$preds2 <- preds2

# 2) WHR is regressed on the predicted values for  BMI + control
# variables
test2 <- paste("whr ~  preds2 + ", paste(pcas, "age.at + sex", sep="",
                              collapse=""), sep="", collapse="")
obj2 <- lm(test2, data=massive.mult)

#3a) The residuals are obtained and then regressed on all instruments
# + control varialbes.
res2 <- residuals(obj2)
massive.mult$res2 <- res2
test3 <- paste("res2 ~ ", for.form, sep="", collapse="")
obj3 <- lm(test3, data=massive.mult)
# (3b) The residuals are regressed on all control variables
test5 <- paste("res2 ~ ", paste(pcas, "age.at + sex", sep="",
                                collapse=""), sep="", collapse="")
obj5 <- lm(test5, data=massive.mult)
# The F-statistic for the effect of the instruments in regression (3a)
```

```r
# are obtained
Fstat.WHR <- anova(obj5, obj3)$F[2]

# 4) F statistic is adjusted with a degrees of freedom correction to
# account for the same genetic variants having been used in the
# regressions of steps 1 and 2.
all.vars <- unique(c(colnames(massive.bmi)[9:84],
                     colnames(massive.whr)[9:47]))
Fstat.WHR.adj <- Fstat.WHR * ((length(all.vars)-1)/length(all.vars))


# Conditional F statistic BMI

# 1) WHR is regressed on full set of genetic instruments +
# control variables, predicted values are obtained
test1 <- paste("whr ~ ", for.form, sep="", collapse="")
obj1 <- lm(test1, data=massive.mult)
preds1 <- fitted.values(obj1)
massive.mult$preds1 <- preds1

# 2) BMI is regressed on the predicted values for  WHR +
# control variables
test2b <- paste("bmi ~  preds1 + ", paste(pcas, "age.at + sex",
                    sep="", collapse=""), sep="", collapse="")
obj2b <- lm(test2b, data=massive.mult)

#3a) The residuals are obtained and then regressed on all
# instruments + control variables
res2b <- residuals(obj2b)
massive.mult$res2b <- res2b
test3b <- paste("res2b ~ ", for.form, sep="", collapse="")
obj3b <- lm(test3b, data=massive.mult)
# (3b) The residuals are regressed on all control variables
test5b <- paste("res2b ~ ", paste(pcas, "age.at + sex", sep="",
                          collapse=""), sep="", collapse="")
obj5b <- lm(test5b, data=massive.mult)
# The F-statistic for the effect of the instruments in regression
# (3a) are obtained
Fstat.BMI <- anova(obj5b, obj3b)$F[2]

# 4) F statistic is adjusted with a degrees of freedom correction
# to account for the same genetic variants having been used in the
# regressions of steps 1 and 2.
Fstat.BMI.adj <- Fstat.BMI * ((length(all.vars)-1)/length(all.vars))
```

#$# Two-sample multivariable MR analysis

The mvmr function (from the MVMR package) is used to obtain the multivariable summary MR estimates for BMI and WHR. In order to employ this function, the summary statistics need to be formatted appropriately. The exposure coefficients, SEs, etc. can be obtained from within the package by extracting them using the mv_extract_exposures() function from the TwoSampleMR function, which will pull the SNPs from the relevant studies and automatically clump them with an LD threshold of 0.001. As our SNP selection is restricted to those identified in individuals of European descent, all that remains is to select these from the exposure object (exposure_dat2) Creating an appropriate object for the outcome needs to be done manually, as the estimates are obtained from individual level UK biobank data. Gene-outcome associations, SEs and P-values were previously obtained for the univariable summary MR analyses and saved in the twosamplEXP objects. These objects are reformatted to the out_dat_tot object, which is then reduced to contain only the relevant SNPs (out_dat). Exposure_dat2 and out_dat are formatted to a single object (using the mv_harmonise_data function). This object can easily be converted to one suitable for the mvmr function, which is used to obtain the relevant IV estimates on the log scale (exponentiated to obtain the rate coefficients reported in the article). The mvmr function from the MVMR package was used in favour of the mv_multiple function of the TwoSampleMR package as the former is more up to date and will in future be integrated into the TwoSampleMR package.

```
# objects that contain the outcome coefficients, SEs and P values. as well as
# effect alleles and effect allele frequencies for BMI and WHR, respectively
load(file="twosampBMI")
load(file="twosampWHR")

str(BMIbig)
twosampBMI <- cbind(twosampBMI, BMIbig$EA, BMIbig$OA, BMIbig$EAF)
colnames(twosampBMI)[13:15] <- c("EA", "OA", "EAF")
twosampWHR <- cbind(twosampWHR, WHRbig$EA, WHRbig$OA, WHRbig$EAF)
colnames(twosampWHR)[11:13] <- c("EA", "OA", "EAF")
# Creating the outcome object

# obtaining relevant parts of the objects - note that the order of SNPs is
# maintained across the out.matrix.bmi.a/out.matrix.whr.a files and the
# BMIbig/WHRbig files
totalSNP <- unique(c(as.character(twosampBMI$SNP), as.character(twosampWHR$SNP)))
effalls <- c(as.character(twosampBMI$EA),as.character(twosampWHR$EA))[-which(
            duplicated( c(as.character(twosampBMI$SNP),
                          as.character(twosampWHR$SNP)))==TRUE)]
otheralls <- c(as.character(twosampBMI$OA), as.character(twosampWHR$OA))[-which(
            duplicated(c(as.character(twosampBMI$SNP),
                         as.character(twosampWHR$SNP)))==TRUE)]
eafs <- c(as.character(twosampBMI$EAF), as.character(twosampWHR$EAF))[-which(
            duplicated(c(as.character(twosampBMI$SNP),
                         as.character(twosampWHR$SNP)))==TRUE)]
outests <- c(twosampBMI[,1], twosampWHR[,1])[-which(duplicated(
                    c(twosampBMI[,1], twosampWHR[,1]))==TRUE)]
outsds <- c(twosampBMI[,2], twosampWHR[,2])[-which(duplicated(
                   c(twosampBMI[,1], twosampWHR[,1]))==TRUE)]
outps <- c(twosampBMI[,4], twosampWHR[,4])[-which(duplicated(
                 c(twosampBMI[,1], twosampWHR[,1]))==TRUE)]

# formatting output to make it suitable for MR function in TwosampleMR package
out_dat_tot <- as.data.frame(cbind(totalSNP, outests, outsds, outps, effalls,
                                   otheralls, eafs, "COUNT", "4", "TRUE"))
colnames(out_dat_tot) <- c("SNP", "beta.outcome", "se.outcome", "pval.outcome",
                           "effect_allele.outcome","other_allele.outcome",
                      "eaf.outcome","outcome", "id.outcome", "mr_keep.outcome")
out_dat_tot$SNP <- as.character(out_dat_tot$SNP)
out_dat_tot$effect_allele.outcome <- as.character(out_dat_tot$effect_allele.outcome)
out_dat_tot$other_allele.outcome <- as.character(out_dat_tot$other_allele.outcome)
out_dat_tot$beta.outcome <- as.numeric(as.character(out_dat_tot$beta.outcome))
out_dat_tot$se.outcome <- as.numeric(as.character(out_dat_tot$se.outcome))
out_dat_tot$pval.outcome <- as.numeric(as.character(out_dat_tot$pval.outcome))
```

```r
out_dat_tot$eaf.outcome <- as.numeric(as.character(out_dat_tot$eaf.outcome))
out_dat_tot$outcome <- as.character(out_dat_tot$outcome)
out_dat_tot$id.outcome <- as.character(out_dat_tot$id.outcome)
out_dat_tot$mr_keep.outcome <- (out_dat_tot$beta.outcome <10)


# Creating the exposure object

# extract exposures using MRbase function, LD clumping is automatically performed
exposure_dat <- mv_extract_exposures(c("ieu-a-73","ieu-a-835"))

# reduce SNPs to the set originally considered (for both the Locke and Shungin
# studies only the European descent SNPs are relevant. The SNPs are given in
# objects BMIbig and WHRbig, previously used for the univariable two-sample MRs)
exposure_dat2 <- exposure_dat[which(exposure_dat$SNP %in%
                              unique(c(as.character(BMIbig$SNP),as.character(WHRbig$SNP)))),]

# reduce outcome data to the SNPs that remain after clumping and after removing
# all SNPs that have not been identified in individuals of European descent
out_dat <- out_dat_tot[which(out_dat_tot$SNP %in% exposure_dat2$SNP),]

# having created the appropriately formatted exposure and outcome objects, the
# data is harmonized (ensuring that the effect direction is consistent across
# exposure and outcome) the harmonization function also formats the two objects
# to one object that can be used in the multivariable two-sample MR function,
# or transformed to be suitable for the mvmr object
both_short <- mv_harmonise_data(exposure_dat2, out_dat)

# obtaining estimates using mv_multiple from TwoSampleMR package
out <- mv_multiple(both_short)


# transforming both_short object for mvmr function from MVMR package
install.packages("remotes")
library(remotes)
install_github("WSpiller/MVMR", build_opts = c("--no-resave-data", "--no-manual"),
               build_vignettes = TRUE)
library(MVMR)


fordata <- format_mvmr(cbind(both_short$exposure_beta[,1],
                            both_short$exposure_beta[,2]),
                            both_short$outcome_beta,
                          cbind(both_short$exposure_se[,1],both_short$exposure_se[,2]),
                          both_short$outcome_se,row.names(both_short$exposure_beta))
head(fordata)

# obtaining estimates using mvmr
outt <- mvmr(fordata)
exp(outt$coef[,1])
exp(outt$coef[,1]-1.96*outt$coef[,2])
exp(outt$coef[,1]+1.96*outt$coef[,2])
```
